# Probing Autism and ADHD subtypes using cortical signatures of the T1w/T2w-ratio and morphometry

**DOI:** 10.1101/2024.08.08.24311239

**Authors:** Linn B. Norbom, Bilal Syed, Rikka Kjelkenes, Jaroslav Rokicki, Antoine Beauchamp, Stener Nerland, Azadeh Kushki, Evdokia Anagnostou, Paul Arnold, Jennifer Crosbie, Elizabeth Kelley, Robert Nicolson, Russell Schachar, Margot J. Taylor, Lars T. Westlye, Christian K. Tamnes, Jason P. Lerch

## Abstract

Autism spectrum disorder (ASD) and attention-deficit/hyperactivity disorder (ADHD) are neurodevelopmental conditions that share genetic etiology and frequently co-occur. Given this comorbidity and well-established clinical heterogeneity, identifying individuals with similar brain signatures may be valuable for predicting clinical outcomes and tailoring treatment strategies. Cortical myelination is a prominent developmental process, and its disruption is a candidate mechanism for both disorders. Yet, no studies have attempted to identify subtypes using T1w/T2w-ratio, a magnetic resonance imaging (MRI) based proxy for intracortical myelin. Moreover, cortical variability arises from numerous biological pathways, and multimodal approaches can integrate cortical metrics into a single network. We analyzed data from 310 individuals aged 2.6-23.6 years, obtained from the Province of Ontario Neurodevelopmental (POND) Network consisting of individuals diagnosed with ASD (n=136), ADHD (n=100), and typically developing (TD) individuals (n=74). We first tested for differences in T1w/T2w-ratio between diagnostic categories and controls. We then performed unimodal (T1w/T2w-ratio) and multimodal (T1w/T2w-ratio, cortical thickness, and surface area) spectral clustering to identify diagnostic-blind subgroups. Linear models revealed no statistically significant case-control differences in T1w/T2w-ratio. Unimodal clustering mostly isolated single individual- or minority clusters, driven by image quality and intensity outliers. Multimodal clustering suggested three distinct subgroups, which transcended diagnostic boundaries, showing separate cortical patterns but similar clinical and cognitive profiles. T1w/T2w-ratio features were the most relevant for demarcation, followed by surface area. While our analysis revealed no significant case-control differences, multimodal clustering incorporating the T1w/T2w-ratio among cortical features holds promise for identifying biologically similar subsets of individuals with neurodevelopmental conditions.

## 1 Introduction

Autism spectrum disorder (ASD) and attention-deficit/hyperactivity disorder (ADHD) are complex and heritable neurodevelopmental conditions that affect about 1.5% and 7% of the population respectively, with higher prevalence in boys (Talantseva et al., 2023; Thomas et al., 2015). Spanning a wide range of function, ASD typically encompasses challenges in social functioning, atypical sensory sensitivity, and stereotyped patterns of movement, interests, and behaviors (American Psychiatric Association, 2022). ADHD is recognized by age-inappropriate levels of inattention, and/or hyperactivity and impulsivity (American Psychiatric Association, 2022). While ASD and ADHD are diagnostically distinct, with different core symptoms, they frequently co-occur. There are for instance reports of children with ADHD showing elevated levels of autistic-like traits (Grzadzinski et al., 2016; Martin et al., 2014), and about 40% of individuals with ASD also presenting with comorbid ADHD (Rong et al., 2021). Moreover, there is considerable overlap in the genetic etiology of ASD and ADHD (Mattheisen et al., 2022). Relatives of individuals with ASD have an increased risk for ADHD, which increases by the level of relatedness (Ghirardi et al., 2018; Jokiranta-Olkoniemi et al., 2016). Consequently, researchers increasingly adopt transdiagnostic approaches, when investigating underlying neural mechanisms of ASD and ADHD.

ASD and ADHD typically manifest in early and mid-childhood, respectively. These are critical periods for brain development, including of the cerebral cortex, which plays a crucial role for cognitive abilities (Gilmore et al., 2018; Norbom et al., 2021). Magnetic resonance imaging (MRI) can indirectly capture these processes, revealing normative decreases in apparent thickness from about year 2, and early increases, and subsequent leveling off, in surface area in typically developing children and adolescents (Bethlehem et al., 2022; Tamnes et al., 2017). Beyond morphometry, a prominent feature of cortical development is a change in its brightness, and variations in cortical intensity can be assessed by computing a ratio between T1-weighted (T1w) and T2-weighted (T2w) scans (Glasser & Van Essen, 2011). The T1w/T2w-ratio has been suggested as a viable MRI based proxy for intracortical myelin (Glasser & Van Essen, 2011; Nakamura et al., 2017; Shafee et al., 2015). This is because cholesterol in myelin is a major determinant for both the T1w and T2w signal, in an inverse manner (Does, 2018; Koenig, 1991; Koenig et al., 1990).

Intracortical myelination is a crucial developmental process providing efficient signal transmission between neurons, and structural support (D. J. Miller et al., 2012; Wake et al., 2011). Multiple studies demonstrate an age-related increase in T1w/T2w-ratio throughout childhood and adolescence, and support the effectiveness of the ratio in mapping global and regional cortical maturational patterns (Baum et al., 2021; Grydeland et al., 2013, 2019; Norbom, Rokicki, Alnæs, et al., 2020; Vandewouw et al., 2019). Oligodendrocyte dysregulation and myelination disruptions have been listed among the putative neurobiological mechanisms for ASD and ADHD (Patterson, 2011; Phan et al., 2020; Richetto et al., 2016) and may therefore be relevant for some individuals who have been diagnosed with these conditions.

As neurodevelopmental conditions, both ASD and ADHD are thought to involve atypical pre- and post-natal brain development. MRI studies have revealed atypical cortical- and white matter microstructure in ASD and ADHD (Ameis et al., 2016; Bezgin et al., 2018; Connaughton et al., 2022; Gharehgazlou et al., 2022; MRC AIMS Consortium et al., 2018; Ohta et al., 2020). Still, previous studies disagree on the presence and direction of structural differences, regional specificity, and the imaging metric most sensitive to these aspects (Hong et al., 2020; Pagnozzi et al., 2018). These discrepancies may in part be attributed to the predominant use of case-control designs. While suitable for homogeneous clinical populations, these designs are subpar when applied to complex conditions like ASD and ADHD as they may dilute their well-recognized phenotypic and etiological heterogeneity (Zabihi et al., 2020).

Recognizing this notion, subtyping ASD and ADHD based on brain imaging could not only clarify previous discrepancies but may shed light on variance in severity, illness progression, treatment response, etiology, and potentially be informative for the development of tailored treatments. Brain-based subtyping has found subcategories both within and across neurodevelopmental conditions (Itahashi et al., 2020; Jacobs et al., 2021; Kushki et al., 2021; Sadat-Nejad et al., 2023; Vandewouw et al., 2023), including structure-based clustering of ASD (Brucar et al., 2023; Hong et al., 2020; Zabihi et al., 2020). Although subtyping based on cortical structure shows potential, studies have yet to identify subtypes based on T1w/T2w-ratio.

Another limitation with the previous literature is that most studies assess a single or a few selected brain metrics separately. This overlooks the probable scenario of cortical variability arising from multiple and partly independent biological pathways, that are indirectly captured best by different imaging metrics. Multimodal approaches can effectively integrate several cortical sources of variability by fusing their similarity matrices into a single similarity network, capturing shared and complementary information and higher-order interactions (Wang et al., 2014). Such approaches could lead to both enhanced parsing and improved neurobiological interpretation.

A final consideration is that, due to convention and availability, many cortical region of interest (ROI) studies default to using the Desikan-Killiany or the Destrieux atlas (Desikan et al., 2006; Destrieux et al., 2010). These atlases are based on gyral folding and anatomical landmarks rather than cyto- or myeloarchitecture which are the main proposed neurobiological drivers of cortical thickness, surface area and T1w/T2w-ratio. A parcellation based on multimodal imaging, including relative myelin content, such as the “multimodal Glasser atlas” (Glasser et al., 2016), could in these cases be more appropriate.

To this end we used, post quality-control, data from 310 children, adolescents, and young adults (89 females) aged 2.6-23.6 years, obtained from the Province of Ontario Neurodevelopmental (POND) Network. Our sample consisted of individuals diagnosed with ASD (n=136), ADHD (n=100), and typically developing (TD) individuals (n=74). Separately for ASD and ADHD, we first employed a case-control design, testing for group differences in cortical microstructure as assessed by the T1w/T2w-ratio in the multimodal Glasser atlas. We then used a spectral clustering approach to find diagnostic-blind subgroups based on the similarity of their regional T1w/T2w-ratio profiles. Finally, we fused Glasser-ROI based T1w/T2w-ratio, cortical thickness, and surface area before using the same clustering approach. Due to clinical heterogeneity, and the likelihood of small effect sizes, if present (Meyer et al., 2001; K. L. Miller et al., 2016), we did not expect statistically significant case-control differences in microstructure between the neurodevelopmental conditions and TD participants. Next, we hypothesized that our clustering approach based on T1w/T2w-ratio would identify transdiagnostic subgroups with relevant differences in social functioning, repetitive behaviors, attention, hyperactivity and total IQ scores. Finally, we hypothesized that incorporating T1w/T2w-ratio into a multimodal clustering approach along with other cortical structural imaging metrics would enhance the ability to group individuals based on shared characteristics.

## 2 Materials and Methods

### 2.1 Participants

The data were acquired from the POND Network, which is collected across four Canadian cites (https://pond-network.ca/). The sample comprises over 3000 individuals spanning an age range from young childhood to young adulthood. Most of the participants have neurodevelopmental disorders or conditions including ASD, ADHD, obsessive-compulsive disorder (OCD), Tourette-Rett- or Fragile X-syndrome (Jacobs et al., 2021; Kushki et al., 2021). The POND Network holds comprehensive demographic, genetic, and behavioral information, along with neuroimaging data for a subset of individuals. The clinical cohort used in the current study was recruited from mental health facilities, primarily based on receiving a diagnosis. TD participants were recruited through public advertising and social media, provided that neither they nor their first-degree relatives had a diagnosis of a neurodevelopmental condition. Parental informed consent, as well as child assent when possible, was obtained for all participants and approval of research ethics was obtained by the Hospital for Sick Children, Toronto, Canada (1000012230) (Baribeau et al., 2019; Sadat-Nejad et al., 2023).

The present study focused on a subset of 484 potential participants recruited between 2016 and 2022 who had both T1w and T2w MRI sequences acquired at a single site, as this site was the only one to obtain sufficient number of images with a voxel resolution of 0.8mm isotropic at the time of the data request for this paper. 117 individuals did not pass MRI based quality control or processing (described below) and another 57 were excluded as they had neurodevelopmental conditions other than ASD or ADHD or missing demographic information. This resulted in a final sample size of 310 participants (89 females) aged 2.6-23.6 years (mean =12.4, SD =4.4). The sample included individuals diagnosed with ASD (n=136), ADHD (n=100), and TD individuals (n=74) (Figure 1).

**Figure 1.**
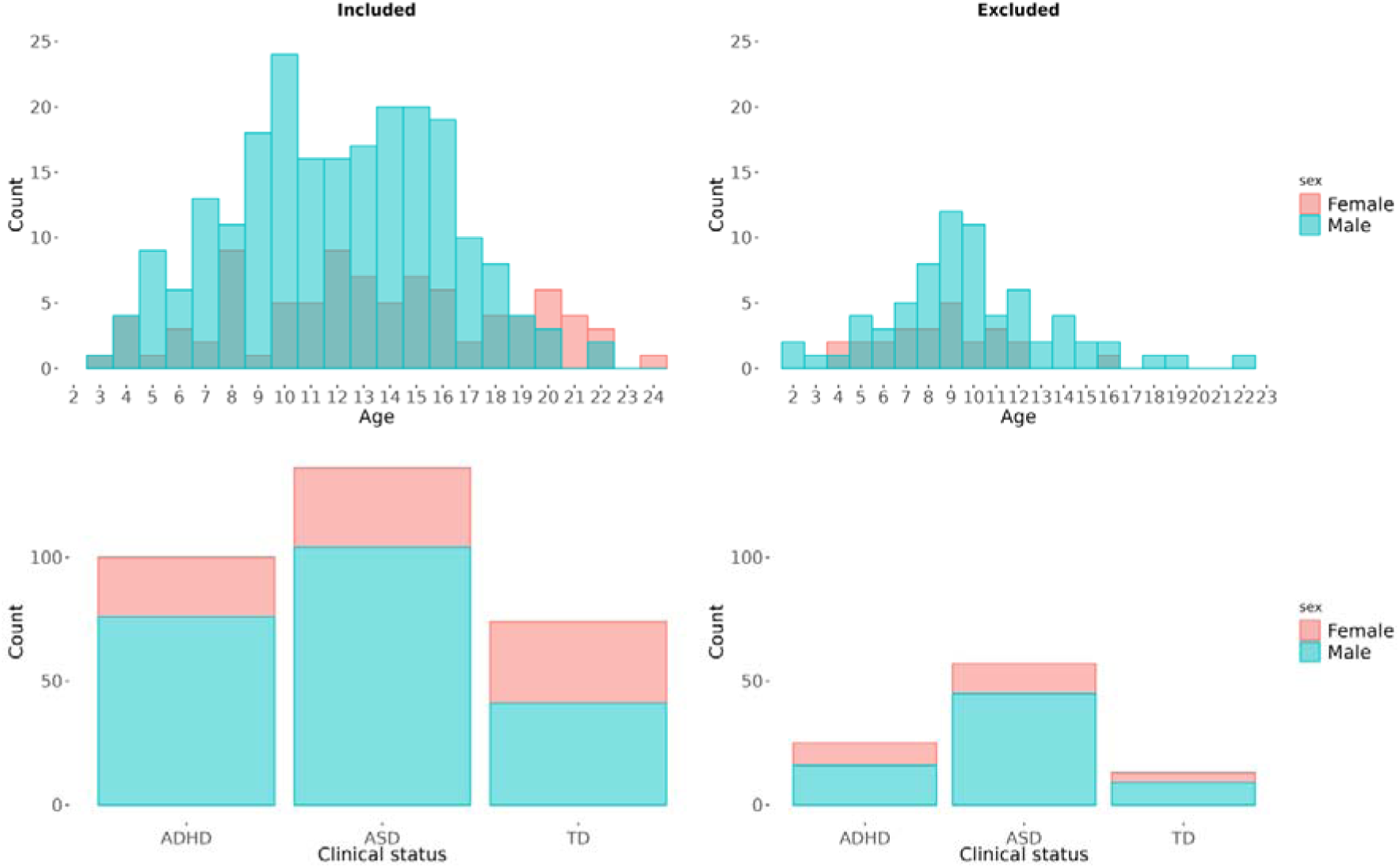
Age, sex and diagnostic histograms. The figure shows the distributions of age, sex, and diagnoses for the final sample (n=310) on the left side, and for the individuals excluded during MRI-based quality control and processing (n=117) on the right side. ASD= autism spectrum disorder, ADHD= attention-deficit/hyperactivity disorder, and TD= typically developing individuals.

### 2.2 Diagnostic-, clinical-, and cognitive measures

Clinical diagnoses were confirmed using the Autism Diagnostic Observation Schedule – 2 (Lord et al., 2012) and the Autism Diagnostic Interview – Revised (Lord et al., 1994) for ASD, and The Parent Interview for Child Symptoms (Ickowicz et al., 2006) for ADHD.

Social functioning was assessed using the Social Communication Questionnaire (SCQ) (Lord & Rutter, 2003), a widely used caregiver report comprising 40 binary items that quantify the presence or absence of non-typical social abilities and behaviors. For our analyses, we utilized the total SCQ score, with higher scores indicating greater social communication deficits. We used the Repetitive Behaviors Scale – Revised (RBS-R) (Bodfish et al., 2000) to assess a broad range of repetitive behaviors including stereotypic, self-injurious and compulsive conduct. The scale consists of 43 items scored by a caregiver on a 0-3 Likert scale with 0 and 3 indicating no and severe presence, respectively. We utilized the total RBS-R score, with higher scores indicating having more repetitive behaviors. Inattentive and hyperactive-impulsive traits were assessed by the Strengths and Weaknesses of ADHD-symptoms and Normal-behavior (SWAN) questionnaire (Swanson et al., 2012), which employs a Likert scale with varying levels across items. Inattention was determined by summing the first 9 items, and the hyperactivity-impulsivity metric by summing items 10-18, with higher scores indicating greater severity. Individuals with missing clinical scores, 72 for SCQ, 75 for RBS-R, and 79 for SWAN, were excluded from the clinical analyses.

Cognitive abilities were assessed with the age-appropriate Wechsler or Stanford– Binet test (Wechsler, David, 2011, 2014). These tests assess multiple dimensions of intelligence, including verbal comprehension, perceptual reasoning, working memory, and processing speed. In the current study we used the age standardized Full-Scale IQ score as a reflection of general cognitive abilities. 69 individuals had missing IQ scores and were excluded from the cognitive analyses.

### 2.3 MRI acquisition, quality control, and preprocessing

MRI data were obtained at The Hospital for Sick Children (SickKids) in Toronto, using a single 3T Siemens MAGNETOM PRISMA scanner. Anatomical scans were acquired using a sagittal T1w MPRAGE sequence, and a sagittal T2w 3D-SPACE sequence. The resolutions for both sequences were 0.8 mm isotropic. Detailed information regarding acquisition parameters is given in the Supplementary Information (SI).

Out of 484 potential participants, 203 individuals had multiple T1w and/or T2w images and/or sessions. To identify the optimal image, a single trained rater (L.B.N) visually inspected the volumes in FSLeyes, giving preference to the earliest session when possible. During this process, 41 individuals were excluded due to inadequate quality of all volumes.

The T1w and T2w images were subsequently processed using a three-step pipeline from the Human Connectome Project (HCP), which is described in detail elsewhere (Glasser et al., 2013). Briefly, step one generates a native space, aligns the T1w and T2w image, applies bias field correction, and creates transformations from native to standard MNI space. Step two, based on FreeSurfer (Fischl, 2012) includes reconstructions of the “white” and “pial” surface, representing the boundaries between grey and white matter, and grey and cerebrospinal fluid, respectively (Dale et al., 1999; Fischl et al., 1999). Cortical thickness is computed based on the vertex-wise distances between these surfaces. Finally, step three maps individual surfaces to a common “fs_LR” space, surface area is calculated by summing the triangle areas converging at each particular vertex of the midthickness surface, and T1w/T2w-ratio maps are created. More specifically, based on methods from Glasser and Van Essen (2011) the T1w volume is divided by the T2w volume, the cortical ribbon voxels are isolated, and voxels suspected of being highly affected by partial voluming are removed. The remaining voxels are mapped onto a midthickness-surface and averaged to produce a single value per vertex. 12 individuals were excluded due to exit-errors or a failure to complete the pipeline within a reasonable timeframe (30 hours).

After image processing, a report was created consisting of screenshots depicting all T1w/T2w-ratio maps. These maps were visually inspected by the same trained rater, who excluded 64 individuals due to inadequate quality. T1w/T2w-ratio, cortical thickness, and surface area were in fs_LR space and with a vertex-wise resolution of 32k. Finally, we parcellated our data using the “Glasser multimodal atlas” (Glasser et al., 2016) obtained from BALSA (https://balsa.wustl.edu/file/3VLx), resulting in 360 cross-hemispheric regions per metric, totaling 1080 features.

As a supplementary analysis we retained only 43% of the full imaging sample, based on additional MRI quality control (QC) steps, including the FreeSurfer output “total number of surface holes” similar to the Euler Number, as described in SI.

### 2.4 Statistical analysis

Based on our sample size, we first conducted a power analysis to estimate the probability that a test would detect true case-control effects (1-beta) and the probability of Type II errors (beta). We utilized the “pwr.t2n.test” function in R and assessed small (Cohen’s d = 0.2) and medium (Cohen’s d = 0.5) effect size detection, using a two-sample t-test with unequal sample sizes, and an alpha level of 0.05. This gave base-estimates and were not adjusted for later multiple comparisons, which lower power further. Calculations were performed separately for ASD and ADHD.

We then tested for group level differences in T1w/T2w-ratio between the clinical groups and TD participants in separate linear models using the Permutation Analysis of Linear Models (PALM) toolbox (Winkler et al., 2014). T1w/T2w-ratio ROIs were added as dependent variables, while dummy coded diagnoses was added as an independent variable. Z-standardized age and sex were included as covariates in the analysis. This resulted in 360 ASD related-, and another 360 ADHD related tests.

To assess statistical significance robustly with a limited sample size, we employed data shuffling with 10,000 permutations. We adjusted for multiple comparisons and contrasts using false discovery rate (FDR) with threshold-free cluster enhancement (Smith & Nichols, 2009) and a significance threshold of 0.05. In a supplementary analysis, we performed identical tests on a subset of the data after more stringent QC of the MRI data (n=209).

The statistical code used in the present paper is available online (https://osf.io/dfvk9/).

### 2.5 Spectral clustering

To mitigate age effects which if unhandled would dominate clustering solutions, we employed normative modelling to age-residualize all 1080 ROIs. For this, we employed Gaussian Process Regression in the PCNtoolkit (Rutherford et al., 2022). First, we estimated ROI-specific normative models as a function of age and sex considering only TD participants, with 5-fold cross validation. We subsequently applied the normative models to our clinical population, allowing us to quantify deviations from the norm through an individual- and ROI-specific Z-score.

We performed spectral clustering in R using the SNFtool (Wang et al., 2014). Initially we tested a range of parameters needed for similarity matrix creation, including alpha levels (0.3-0.8) and k-nearest-neighbors (KNN) (11-31). To guide our selection, we evaluated cluster solutions spanning 2 to 10 clusters using the Dunn Index and Silhouette Score, as these metrics can operate directly on the distance matrix. Based on these assessments, we selected an alpha level of 0.3, and a KNN value of 23.

For unimodal clustering we incorporated the 360 T1w/T2w-ratio ROI z-scores from individuals with ASD and ADHD only (n=236) as features. From these, we calculated the pairwise Euclidean distances between individuals. This matrix was then converted into a similarity matrix, where a mathematical transformation emphasizes closer relationships over distant ones while ensuring that similarities are comparable regardless of the density of the data points, as detailed in (Wang et al., 2014). Spectral clustering was then applied to the similarity matrix, exploring solutions of 2 to 10 with 100 iterations.

Similarly, for multimodal clustering, normative modelling-based z-scores from T1w/T2w-ratio, cortical thickness, and surface area were used as features to create three separate distance matrices. They were then converted into three similarity matrices. We then utilized a similarity network fusion technique to combine the distinct matrices into a unified similarity network. This approach minimizes signal dilution which might be associated with other concatenation methods, preserving the integrity of individual cortical sources (Wang et al., 2014). Spectral clustering was then performed on the fused similarity matrix, testing cluster solutions from 2 to 10 with 100 iterations. To test whether the demarcations were influenced by residual age effects, we also performed multimodal clustering on subsets of individuals with narrower age ranges (SI Methods, SI Figure 1).

To identify the optimal number of clustering solutions, we used the “eigengap” and “rotation cost” (Wang et al., 2014). Eigenvalues, obtained from the Laplacian matrix offer valuable insights into the connectivity structure of the data. A large difference between eigenvalues, or a large eigengap, suggests the presence of well-separated clusters. Rotation cost is derived from constructing a rotation matrix using the eigenvectors obtained from the Laplacian matrix. To encourage that each datapoint is predominantly assigned to a single cluster, a sparsity constraint (each row having at most one non-zero entry) was introduced. The optimal number of clusters can be decided by the configuration that achieves minimal rotation cost, indicating that the rotation matrix aligns the eigenvectors in a manner that adheres to the sparsity constraint. When estimators diverged, emphasis was put on rotation cost, as it has been reported to be more stable (Wang et al., 2014).

For optimal cluster solutions we tested whether sub-groups showed statistically significant differences in cognitive or clinical scores. Shapiro-Wilk-, and Levene’s tests indicated that assumptions for parametric testing were violated, due to skewed distributions of most sub-groups, and heteroscedastic inattentive scores. We therefore performed non-parametric testing, i.e. quantile regression in R targeting the median. P-values were estimated using bootstrap methods with 1000 repetitions. For each model, cognitive and clinical scores were included as dependent variables, while cluster membership was incorporated as a categorical independent variable. Age was included as a covariate as we observed modest correlations between age and several of the dependent variables. Sex showed negligible associations with the dependent variables and was therefore not included as a covariate. P-values were adjusted for multiple comparisons by FDR using Benjamini-Hochberg’s procedure and a significance threshold of 0.05. Each tested clustering solution was treated as a separate family.

To identify cortical imaging features that were particularly important for subgrouping, we employed Normalized Mutual Information (NMI). This measure quantifies the similarity between subgroups formed when individuals are clustered on a single feature with those from the actual fused network. A score of 1 indicates perfect agreement, meaning that the individual feature produces identical groupings, while a score of 0 signifies no agreement (Wang et al., 2014). We ranked cortical features by their NMI scores, highlighting the top 10% as particularly relevant.

## 3 Results

### 3.1 Case-control comparisons

Power analyses indicated that we had a high chance of detecting true differences with medium effect sizes in cortical microstructure between TD participants and individuals with ASD (93%) and ADHD (91%) separately. Correspondingly, the chances of committing a type II error were 7% and 9% for ASD and ADHD assessments, respectively. Regarding the detection of small differences, our analyses had limited statistical power, with a 28% chance of detecting such differences within our ASD comparisons, and 26% for ADHD. Correspondingly, the likelihood of committing a type II error were 72% and 74%, respectively.

Permutation testing revealed no significant group differences in T1w/T2w-ratio between individuals with ASD and TD, or individuals with ADHD and TD. Our supplementary analyses, performing the same assessments on a subset with a more stringent QC, also yielded no group differences. Cortical maps showing Cohen’s d values, masked by *uncorrected* p-values thresholded at a minimum of 0.05 (−log(p) = 1.3) are presented in SI Figure 2.

### 3.2 Unimodal clustering

Normative modelling successfully minimized the associations between the T1w/T2w-ratio features and age as presented in SI Figure 3.

Unimodal clustering of T1w/T2w-ratio features from individuals with neurodevelopmental conditions exclusively showed one relatively large cluster (which further divided with more fine-grained parsing), with remaining groupings consisting of single-individual clusters, or clusters with a very limited number of individuals (Figure 2, SI Figure 4). Estimators revealed that a 4-subgroup partition (eigengap= 0.194, rotation cost=177.314), closely followed by a 3-subgroups partition (eigengap= 0.197, rotation cost= 186.563), were the most optimal solutions. Further investigations into these clusters revealed a similar distribution of diagnoses (SI Figure 5). However, individuals appeared to be singled out based on either showing a highly local increase in T1w/T2w-ratio, making them an outlier on a single feature, or showing globally increased T1w/T2w-ratio (SI Figure 6), also coupled with a high number of surface holes (SI Figure 7). T1w/T2w-ratio features that were of particular relevance for parsing are presented in SI Figure 8. Estimators for the cluster solutions are presented in SI Figure 9.

**Figure 2.**
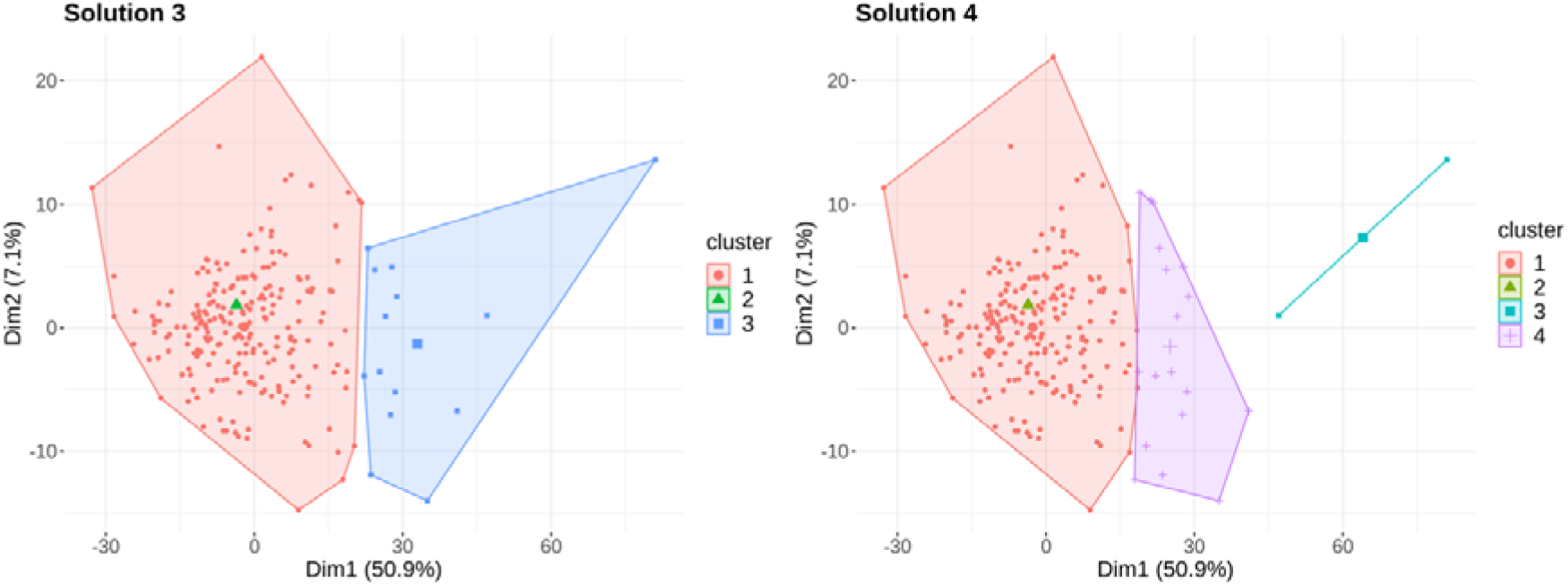
Unimodal clustering solutions. The figure illustrates cluster solutions 3 and 4 from clustering based on T1w/T2w-ratio features. Individuals are plotted according to the first two principal components (Dim1 and Dim2), that explained the majority of the variance in the data. Cluster centroids are depicted as slightly larger shapes.

### 3.3 Multimodal clustering

As for the T1w/T2w-ratio features, normative modelling successfully minimized the associations between cortical thickness features and age. Surface area features showed similar associations, which were negligible from the outset, as presented in SI Figure 3.

Multimodal clustering of all 1080 features from T1w/T2w-ratio, cortical thickness and surface area resulted in groups of similar size (Figure 3, SI Figure 10). The distributions of age, sex, clinical diagnosis, number of surface holes, and brain volumes across all solutions are presented in SI Figures 11-15. The top 10% of cortical features particularly relevant for distinguishing individuals, were T1w/T2w-ratio features, followed by surface area features (Figure 4). The T1w/T2w-ratio features were symmetrically spread across hemispheres and primarily located within lateral and medial parietal regions, extending into occipital regions, and the pre-central gyrus. Particularly relevant surface area regions were also located symmetrically across hemispheres and primarily within lateral temporal- and medial frontal regions. Heatmaps of individual cortical feature scores, segmented by subgroup, generally showed global differences in cortical structure across groups.

**Figure 3.**
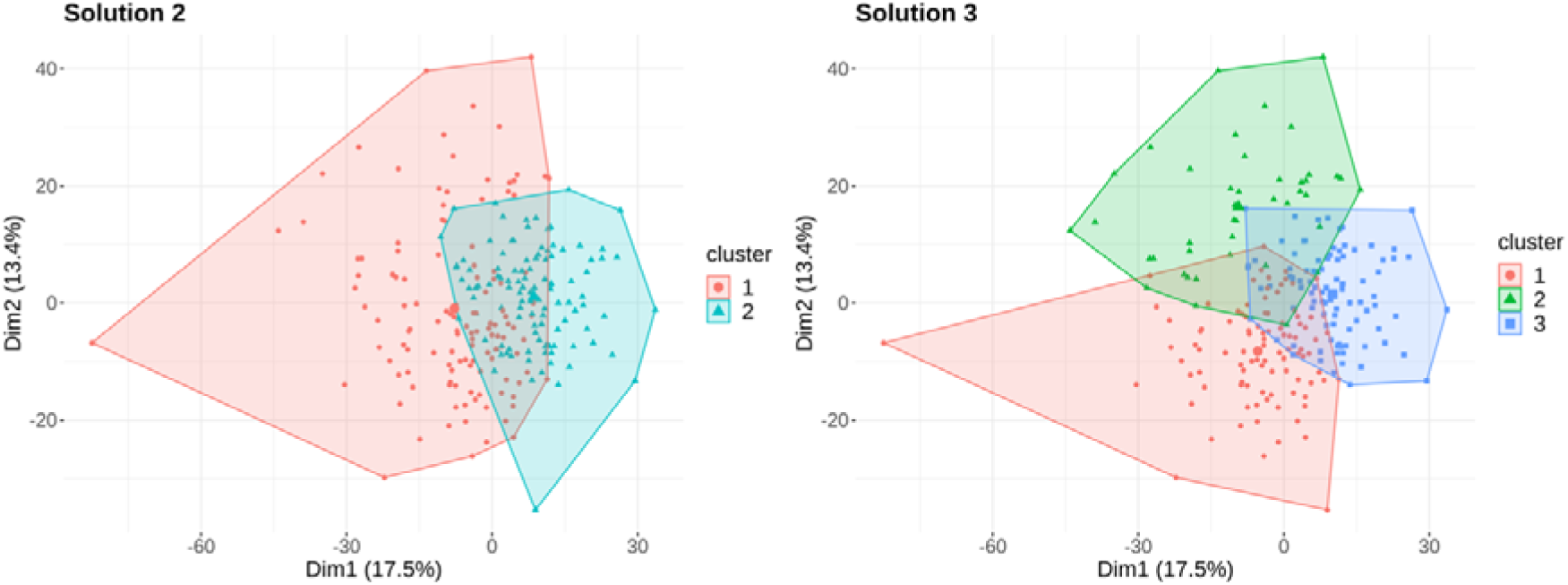
Multimodal clustering solutions. The figure illustrates cluster solutions 2 and 3 from multimodal clustering based on T1w/T2w-ratio, cortical thickness and surface area. Individuals are plotted according to the first two principal components (Dim1 and Dim2), which explained most of the variance in the data. Cluster centroids are depicted as slightly larger shapes.

**Figure 4.**
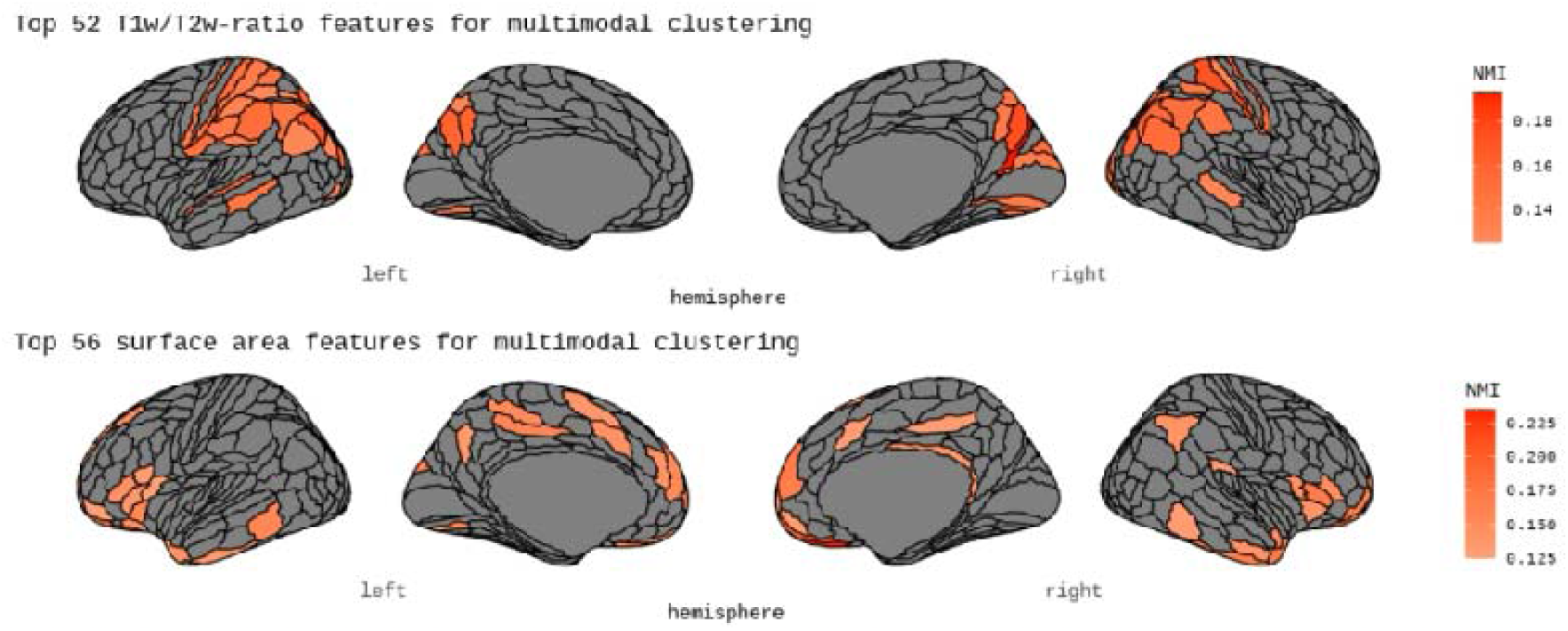
The top 10% most central cortical features for multimodal clustering. The figure displays the top 10% most central cortical features for clustering, based on the highest ranked normalized mutual information (NMI).

Estimators revealed that partitioning into 3 subgroups (eigengap= 0.045, rotation cost= 199.812) yielded the most optimal solution, which was followed by partitioning into 2 subgroups (eigengap= 0.036, rotation cost= 209.348). Estimators for all tested cluster solutions are presented in SI Figure 16. The three-group parsing appeared proportionate on sex, diagnostic categories, and number of surface holes. Individuals within Cluster 1 appeared to have a slightly higher median age compared to those in Cluster 3, while individuals in Cluster 2 appeared to exhibit lower total brain volume - excluding ventricle volume, compared to the other clusters (Figure 5). Individuals within Cluster 1 exhibited slightly higher T1w/T2w-ratio, thinner cortex, and somewhat larger surface area. Individuals within Cluster 2 exhibited higher ratio coupled with much smaller surface area, while individuals within Cluster 3 generally displayed the reverse pattern compared to those in Cluster 1 (SI Figure 17). Cortical patterns remained largely consistent when assessing sub-groups with narrower age ranges (SI Results, SI Figure 18-20).

**Figure 5.**
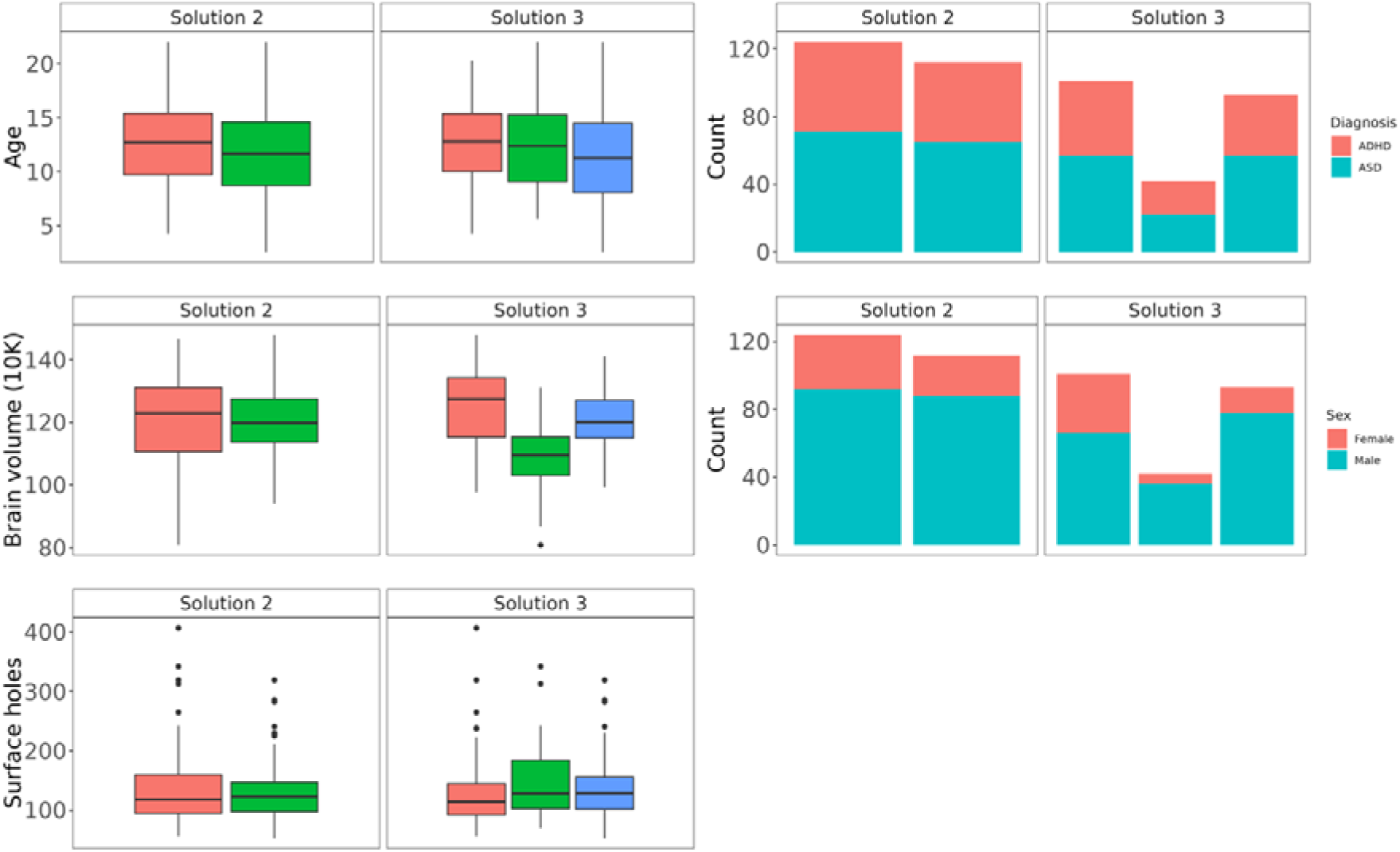
Distribution of age, diagnosis, brain volume, sex, and number of surface holes for multimodal clustering solutions 2 and 3. The figure shows boxplots illustrating the distribution of age, brain volume scaled by a factor of 10k and the number of surface holes, alongside bar plots showing the proportion of diagnoses and sex across the two solutions.

In the two-subgroup parsing, the first cluster appeared to be a combination of what was previously described as Cluster 1 and 2, whereas the second cluster closely resembled the previously described Cluster 3 (SI Figure 17).

We observed modest correlations between age and several of the dependent variables within our quantile regression: IQ (r = -0.02), Repetitive Behaviors (r = -0.03), SCQ Score (r = 0.14), Inattentiveness (r = -0.22), and Hyperactive-Impulsive Behavior (r = -0.27) and therefore included age as a covariate in these analyses. We found no statistically significant differences between the three subgroups for any of the clinical or cognitive measures. Figure 6 shows the distribution of scores after age-residualization. Uncorrected findings indicated that individuals within Cluster 1 had higher median IQ scores (t= -2.82, p= 0.005, corrected p = 0.075) compared to individuals within Cluster 3 and experienced less inattention (t= 2.12 p = 0.036 corrected p= 0.18) and engaged in fewer repetitive behaviors (t = 2.12, p = 0.036, corrected p = 0.18) than individuals within Cluster 2. Estimates were corrected across 15 tests. Similar null findings were generally observed among individuals categorized into two subgroups (see distribution in SI Figure 21) with a noticeable exception of individuals within Cluster 1 showing higher IQ scores (t=-2.65, p= 0.009 corrected p= 0.045) compared to individuals within Cluster 2, also after correction, but here across 5 tests. All t-statistics and corresponding p-values are presented in SI Table 1 and 2.

**Figure 6.**
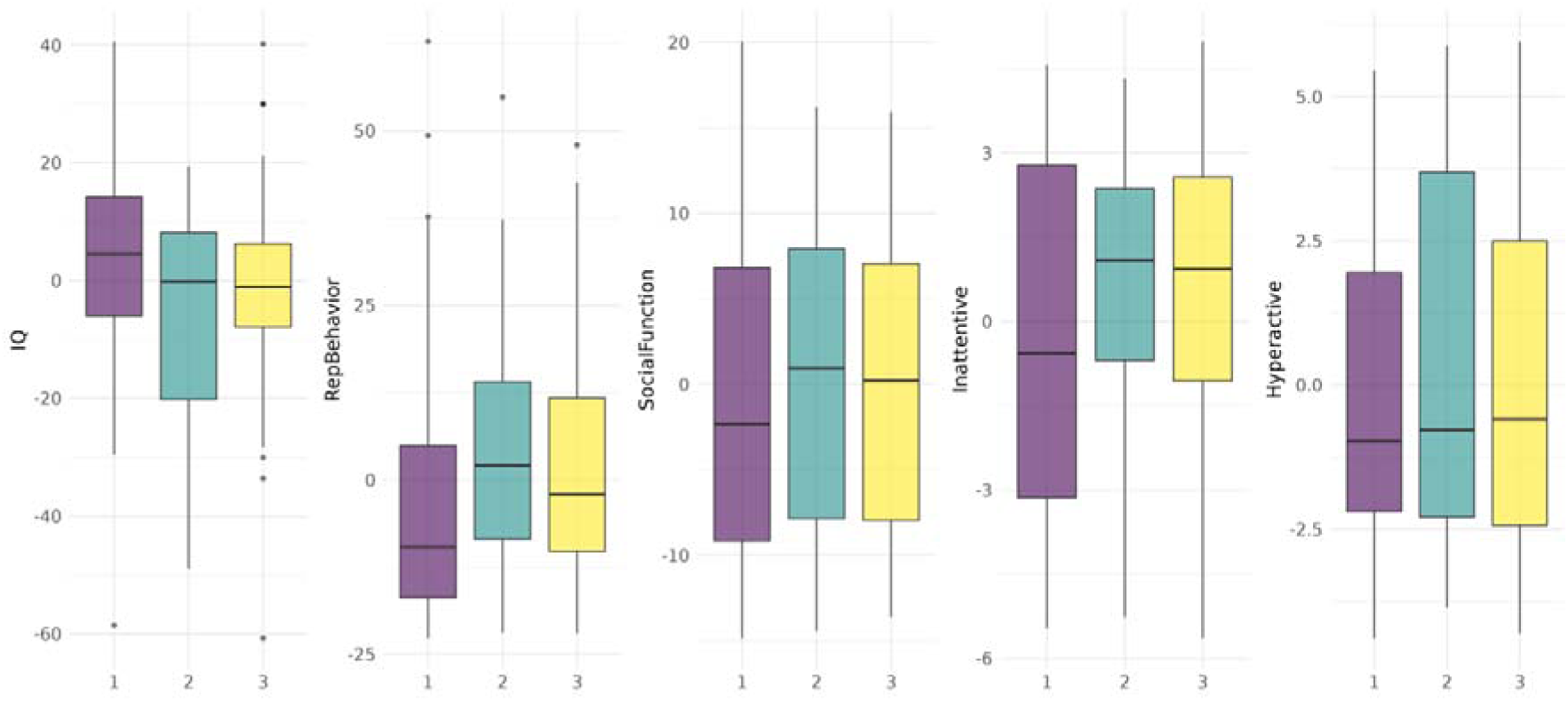
Distribution of clinical and cognitive scores for multimodal clustering solution 3. The figure shows boxplots illustrating the distribution of IQ, repetitive behaviors (RepBehavior), social functioning (SocialFunction), inattentive and hyperactive scores after age-residualization. Individuals with missing scores: IQ n = 69, Social Communication Questionnaire (SCQ) n = 72, Repetitive Behaviors Scale – Revised (RBS-R) n = 75, and Strengths and Weaknesses of ADHD-symptoms and Normal-behavior (SWAN) n = 79 are not included in the plots.

## 4 Discussion

Intracortical myelination is essential for typical brain development, and cortical dysmyelination is one of several candidate mechanisms for neurodevelopmental disorders such as ASD and ADHD (Henriquez-Henriquez et al., 2020; Hettwer et al., 2022; Patterson, 2011; Richetto et al., 2016). In this study, initial case-control comparisons using T1w/T2w-ratio as an intracortical myelin proxy did not reveal consistent differences between diagnostic groups. However, T1w/T2w-ratio features were the most central within multimodal clustering for parsing individuals based on distinct cortical signatures. These subgroups cut across diagnostic categories but did not show different clinical and cognitive profiles.

In line with our hypothesis, we found no differences in cortical microstructure when comparing either participants with ASD or ADHD to their TD peers. Our findings correspond well with a previous smaller-scale study that also found no difference in T1w/T2w-ratio when comparing 21 children with ASD to 16 TD peers, aged 1-5 years (Chen et al., 2022). A few studies have also assessed the related and inverse T1w intensity metric grey/white matter contrast (GWC) in ASD, yielding discrepant findings. One longitudinal study of 20 toddlers reported GWC variations linked to ASD diagnosis (Godel et al., 2021), but the small sample size warrants caution. Another study reported lower regional GWC in 98 adults with ASD compared to 98 TD peers (Andrews et al., 2021), a pattern which a separate youth study (77= ASD, 76=TD) found to be more prominent before 15 years of age (MRC AIMS Consortium et al., 2018). We did not expect case-control differences in T1w/T2w-ratio as dysmyelination represents only one of several candidate etiological mechanisms for ASD and ADHD. Other prominent theories include dysregulation of transcription and translation, synaptic proteins, epigenetics, and immunoinflammatory responses (Jiang et al., 2022; Mehta et al., 2019). Therefore, it is probable that myelin disruptions are implicated for a *subgroup* of youths, and that these effects might be diluted within a case-control design (Zabihi et al., 2020).

Unimodal clustering of T1w/T2w-ratio features resulted in one large group, alongside three smaller groups with single-, or a limited number of individuals. Contrary to our expectations clusters were not distinguished by phenotypically relevant information, but rather by poor image quality and their status as a single-feature or global T1w/T2w-ratio outlier. For this reason, we chose not to pursue further phenotypic analyses based on unimodal clustering. Since no prior studies have assessed clustering based on T1w/T2w-ratio or other intensity metrics, comparing the parsing ability of cortical microstructure is challenging. It is possible that unimodal clustering was dominated by technical variability, while the integration of complementary cortical metrics provided contextual information that highlighted the biologically relevant contributions of the T1w/T2w ratio. Notably, the spatial patterns of the T1w/T2w ratio features that were central for demarcation differed between unimodal and multimodal clustering. Multimodal clustering thus appears to reduce noise and enhance biologically meaningful variation, reinforcing its utility for subgroup identification.

Incorporating microstructure and standard morphometry into a multimodal clustering approach identified three similarly sized trans-diagnostic groups. Within the first cluster, individuals exhibited somewhat brighter and thinner cortex, along with somewhat larger surface area. Despite our effective efforts to minimize age effects prior to clustering, these patterns align with the classic maturational patterns typically observed in MRI-based assessments of cortical development in youth (Norbom et al., 2021). Cortical patterns remained largely consistent, however, in sensitivity analyses, where residual age effects were further mitigated by focusing on subsets of individuals within narrow age ranges.

While inferring neurobiology from the mm-scale of MRI is challenging, such patterns are collectively suggested to reflect increased levels of cortical myelin and axon caliber, as well as dendritic arbor remodeling and reductions in glial cells (Huttenlocher & Dabholkar, 1997; Petanjek et al., 2011; Peter R., 1979; Seldon, 2005; Vidal-Pineiro et al., 2020).

Individuals within the third cluster exhibited cortical patterns contrasting those in cluster one, likely indicating reversed neurobiological interpretations. Participants within the second cluster on the other hand, displayed a brighter cortex combined with markedly smaller surface area. This combination is somewhat unusual from a neurobiological standpoint but could be related to having higher levels of intracortical myelin but an overall smaller brain. Correspondingly, individuals in the second cluster appeared to have smaller total brain volumes compared to the other groups.

Our multimodal clustering findings are somewhat consistent with recent studies that have used partly overlapping samples from the POND Network. One study parsed individuals with ASD, ADHD, OCD and TD participants, based on cortical and subcortical brain structure, and social communication scores. The approach identified three subgroups, with distinct yet transdiagnostic structural signatures and differing levels of social impairment (Kushki et al., 2021). Another study, parsing ASD, ADHD, and TD participants based on the structural co-variance of cortical and subcortical brain structure identified two trans-diagnostic clusters. Clusters differed in age and sex as well as adaptive functioning, inattention, hyperactivity and IQ (Sadat-Nejad et al., 2023). More generally, our findings also align with previous structure-based clustering approaches focusing solely on ASD, which have identified between 2-5 biological subtypes (Brucar et al., 2023; Hong et al., 2020; Zabihi et al., 2020). In a methodologically similar study, researchers initially applied normative modelling to a group of 206 TD individuals before performing cortical thickness-based spectral clustering on 316 individuals with ASD. Their investigation revealed 5 putative subtypes, displaying distinct clinical, behavioral and genetic characteristics (Zabihi et al., 2020).

While Kushki et al., (2021), Sadat-Nejad et al., (2023) and Zabihi et al., (2020) identified clinically relevant clusters, comparisons of our subgroups did not reveal statistically significant differences in clinical or cognitive scores. Due to the stringent requirements of the T1w/T2w metric which required matched high-quality sequences, both of which were at sub-mm isotropic resolution, our study had a smaller sample size than previous studies that performed clustering on standard morphometry. Reflecting a consistent pattern, patients characterized by what might be seen as having a “more mature” cortex (cluster 1) tended to exhibit higher IQs compared to those with a “less mature” cortex (cluster 3), and experience less inattention and engage in fewer repetitive behaviors than those with markedly smaller cortical surface area (cluster 2). A larger sample might be necessary to determine whether these observed uncorrected findings are spurious. However, biologically based subtyping may also identify differences less related to psychopathology and be more reflective of dominant variance in the data (Dinga et al., 2019) such as a similarities in brain size, age, maturational processes, and possibly socioeconomic constructs. One way to increase clinical sensitivity could be to use Canonical Correlation Analysis or similar methods to constrain the search for biologically based subtypes within dimensions of variation specifically related to psychopathology (Alnæs et al., 2020). It is also possible that, despite its importance for brain-based parsing of youths, T1w/T2w-ratio variation, suggestive of variations in intracortical myelin, may not be the primary driver of clinical manifestations observed in atypical neurodevelopment.

As hypothesized, T1w/T2w-ratio features appeared to hold central and complementary information for clustering, and notably, specific features emerged as the most discriminative factors for distinguishing among participants. This was followed by the discriminatory capacity of surface area. This suggests that microstructural variance within neurodevelopmental populations is highly relevant, even after minimizing the effects of age. While no previous studies have performed multimodal clustering with T1w/T2w-ratio, the emphasis on cortical microstructure over morphometry in youth aligns with findings from other multimodal developmental studies. For instance, a study involving almost 2,600 participants aged 3-23 years demonstrated that the related intensity metric GWC, accounted for the majority of cortical variance, surpassing standard morphometry measures. Moreover, GWC showed stronger associations with age compared to standard morphometry (Norbom, Rokicki, Meer, et al., 2020) and socioeconomic diversity, as separately tested in about 10,000 children (Norbom et al., 2024) surpassing the well-established link between socioeconomic diversity and surface area. Our findings add to the body of literature that underscores the sensitivity of intensity metrics in neurodevelopmental research. Future studies aiming to parse neurodevelopmental populations should consider including the T1w/T2w ratio.

There are several limitations to our study. First, radiofrequency transmit field (B1+) correction of T1w/T2w-ratio maps is not yet available in the open-access HCP pipeline, and we therefore did not perform this correction. This bias could correlate with our variables of interest (Glasser et al., 2022; Nerland et al., 2021). Second, while having the benefit of being based on conventional MRI sequences, T1w/T2w-ratio is by no means a straightforward myelin marker. Although it is supported by histological and quantitative relaxometry studies, its links to myelin-related genes and more recognized quantitative proxies are not always consistent (Hagiwara et al., 2018; Righart et al., 2017; Ritchie et al., 2018; Uddin et al., 2019). Third, within psychology, psychiatry (Meyer et al., 2001) and multimodal imaging (Miller et al., 2016) effect sizes are often small. In our study we had limited statistical power to detect small case-control differences if present. Fourth, the goal was not to establish ground truth “biotypes”, but to probe the utility of cortical microstructure in redefining typical and non-typical neurodevelopment, beyond diagnostic categories. Consequently, we did not employ rigorous tests to validate cluster solutions against a null hypothesis (Dinga et al., 2019). Finally, while our findings generally aligned with previous literature, reliability can only be established through replication. There are currently no other datasets with the relevant neurodevelopmental disorders and T1w and T2w sequences with adequate isotropic voxel resolution (< 1mm, preferably 0.7mm), and thus we encourage future investigations into these aspects.

In conclusion, combining cortical macro- and microstructure for parsing of non-typical neurodevelopment revealed distinct subgroups with unique combinations that transcended the diagnostic boundaries of ASD and ADHD. Although microstructural differences were not apparent at the diagnostic group level, T1w/T2w-ratio emerged as a key discriminator for subtyping, followed by surface area. The incorporation of T1w/T2w-ratio in developmental studies offers both a potential for enhancing our understanding of neurodevelopmental diversity and a more precise approach for delineating cortically similar individuals than standard morphometry alone.

## Data Availability

All data produced in the present study are available upon reasonable request to the authors. The statistical code used in the present paper is available online (https://osf.io/dfvk9/).

## Acknowledgments

Data were obtained from the Province of Ontario Neurodevelopmental Network (POND) (https://pond-network.ca/), and with the support of Ontario Brain Institute (PIs: E. Anagnostou, J. Lerch) an independent non-profit corporation, funded partially by the Ontario government. The findings reported in this study are the sole responsibility of the authors.

## Funding

This work was supported by the Research Council of Norway (#223273, #248238, #249795, #276082, #286838, #288083, #323951, # 341355), the South-Eastern Norway Regional Health Authority (#2021070, #2023012, #500189), KG Jebsen Stiftelsen, the ERA-Net Cofund through the ERA PerMed project IMPLEMENT, and the European Research Council under the European Union’s Horizon 2020 research and Innovation program (ERC StG Grant #802998).

## Declaration of Generative AI and AI-assisted technologies in the writing process

During the preparation of this work the corresponding author used chatGPT 4.0 in order to ensure that specific sentences adhered to English language norms and to enhance readability. After using this tool, the author reviewed and edited the content as needed and takes full responsibility for the content of the publication.

## Notes

### Competing Interest Statement

The authors have declared no competing interest.

### Funding Statement

This work was supported by the Research Council of Norway
The South-Eastern Norway Regional Health Authority
The ERA-Net Cofund through the ERA PerMed project IMPLEMENT, and the European Research Council under the European Unions Horizon 2020 research and Innovation program (ERC StG Grant #802998).

### Author Declarations

The ethics committee of the Hospital for Sick Children, Toronto, Canada gave ethical approval for this work (1000012230).

### Summary of Updates

This version includes additional supplemental analyses based on reviewer suggestions

