## Supplementary Information for "Probing Autism and ADHD subtypes using cortical signatures of the T1w/T2w-ratio and morphometry"

**Methods and Materials**

*T1w and T2w acquisition parameters*

MRI data was acquired on a single 3T Siemens MAGNETOM PRISMA scanner. The T1-weigthed (T1w) sequence was a sagittal Magnetization Prepared Rapid Gradient Echo (MPRAGE) scan, with repetition time (TR) = 1870ms, echo time (TE) = 3.10 ms, flip angle = 9°, and voxel resolution = 0.8mm isotropic. The scan had an acceleration factor in the phase-encoding (PE) direction of 2, and an acquisition time (TA) of 5 minutes and 3 seconds. The T2-weigthed (T2w) sequence was a sagittal turbo spin echo (TSE) scan, implemented with the Sampling Perfection with Application optimized Contrast using different flip angle Evolution (SPACE) technique. It had TR = 3200ms, TE= 409ms, and voxel resolution = 0.8mm isotropic. The scan had an acceleration factor in the PE direction of 2, and a TA of 6 minutes and 18 seconds.

*Additional MRI quality control*

We assessed whether null findings were due to variations in data quality, which might obscure true group differences. Mimicking one SD we found the 68% of individuals with the lowest amount of "total surface holes," which is the input for calculating FreeSurfer's Euler number. We then identified participants within two SDs of global z standardized T1w/T2w. 2 SDs were chosen to exclude extreme values that could pertain to image quality while preserving normal variations associated with phenotypes such as age. We intersected these individuals, retaining only those who exhibited both the fewest surface holes and were within SDs of global T1w/T2w-ratio values. The final sensitivity-sample consisted of 209 individuals, which corresponds to 43% of the full MRI sample (n=484) and 57% of the originally quality control (QC)’d MRI sample (n=367). The sensitivity-sample consisted of 87 individuals with autism spectrum disorder (ASD), 70 individuals with attention-deficit/hyperactivity disorder (ADHD), and 52 typically developing (TD) individuals.

*Residual Age-Effect Sensitivity Analysis*

To address concerns that age could influence clustering demarcation despite initial age residualization via normative modeling, we re-ran the clustering on a subset of the sample with a narrower age range. Specifically, we first averaged the age-residualized ROIs to derive global metrics (e.g., mean cortical thickness, total surface area, and mean T1w/T2w ratio) and Z-standardized these for plotting purposes. We then plotted these metrics against the patients' ages to identify periods of minimal variation, which guided the selection of a narrower age range and a more focused sample (Supplementary Figure 1).

We subsequently re-ran the multimodal clustering analysis using the original parameters but restricted it to two subgroups, and only for what was originally deemed the optimal cluster solution. The first subgroup (n = 180) was defined by the black-to-black dotted lines from the age-stability assessment (Supplementary Figure 1), including individuals aged 8–18 years. The second subgroup applied a stricter cut-off (n = 127), corresponding to individuals aged 8–15 years, as defined by the black-to-grey dotted lines.

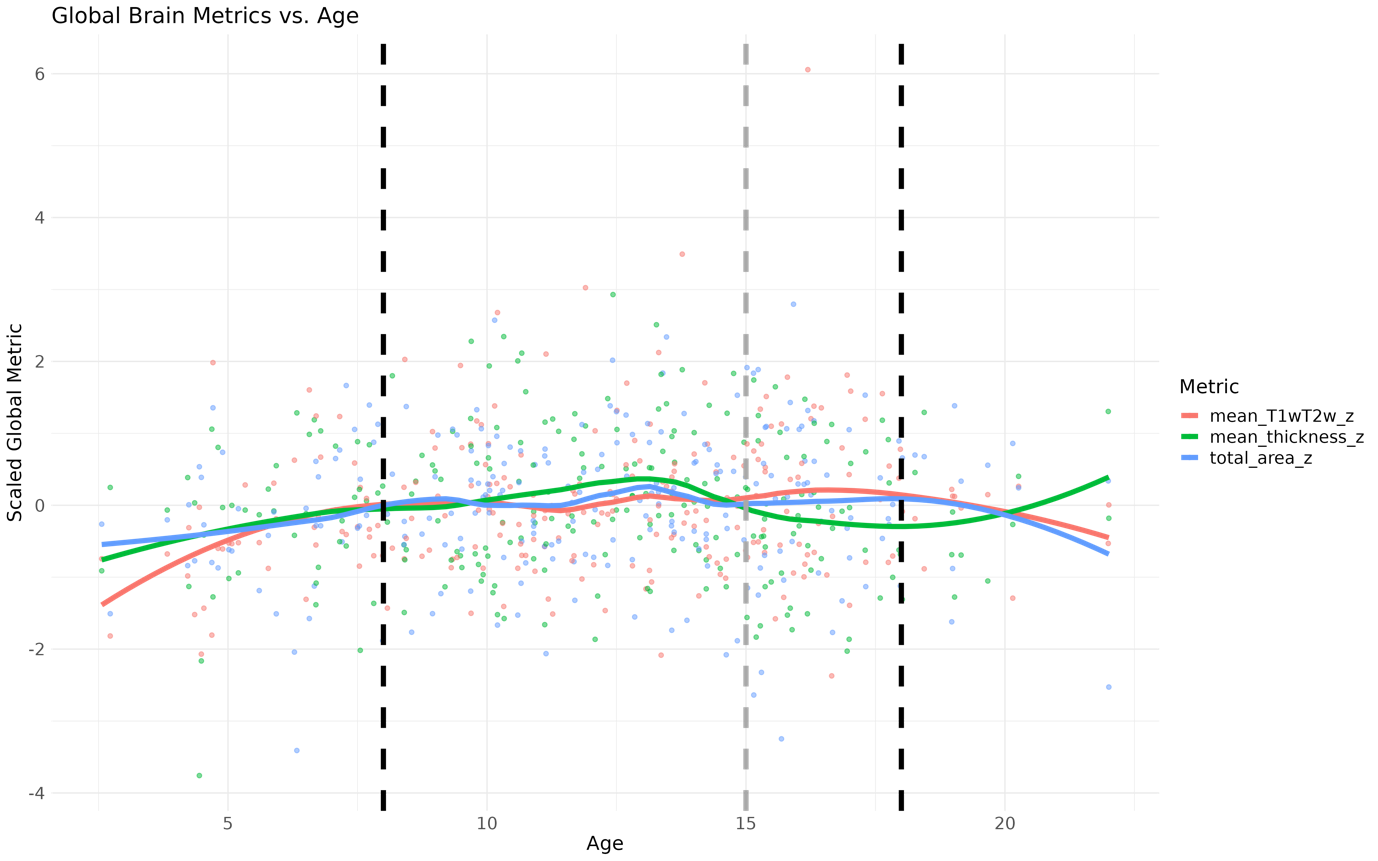

Supplementary Figure 1. The figure illustrates z-scores of age residualized global cortical metrics (mean cortical thickness, total surface area, and mean T1w/T2w ratio) plotted across the age range of the patients. Each point represents an individual, and the colored lines depict smoothed trends for each metric. The black dotted lines indicate the range (8-18 years) where the metrics showed relatively less age-related differences, while the grey dotted line representing an even stricter cut-off (8-15 years).

**Results**

*Case-control assessments*

Permutation revealed no significant group differences in T1w/T2w-ratio when comparing ASD-, or ADHD to TD peers. Likewise, our supplementary analyses, performing the same assessments on the reduced sensitivity-sample, based on additional MRI QC steps, yielded no statistically significant group differences. This indicates that null findings are not caused by variations in imaging quality. Cortical maps of Cohen’s d distributions, masked by *uncorrected* p-values thresholded at a minimum –logp of 1.3 are presented in Supplementary Information (SI) Figure 2.

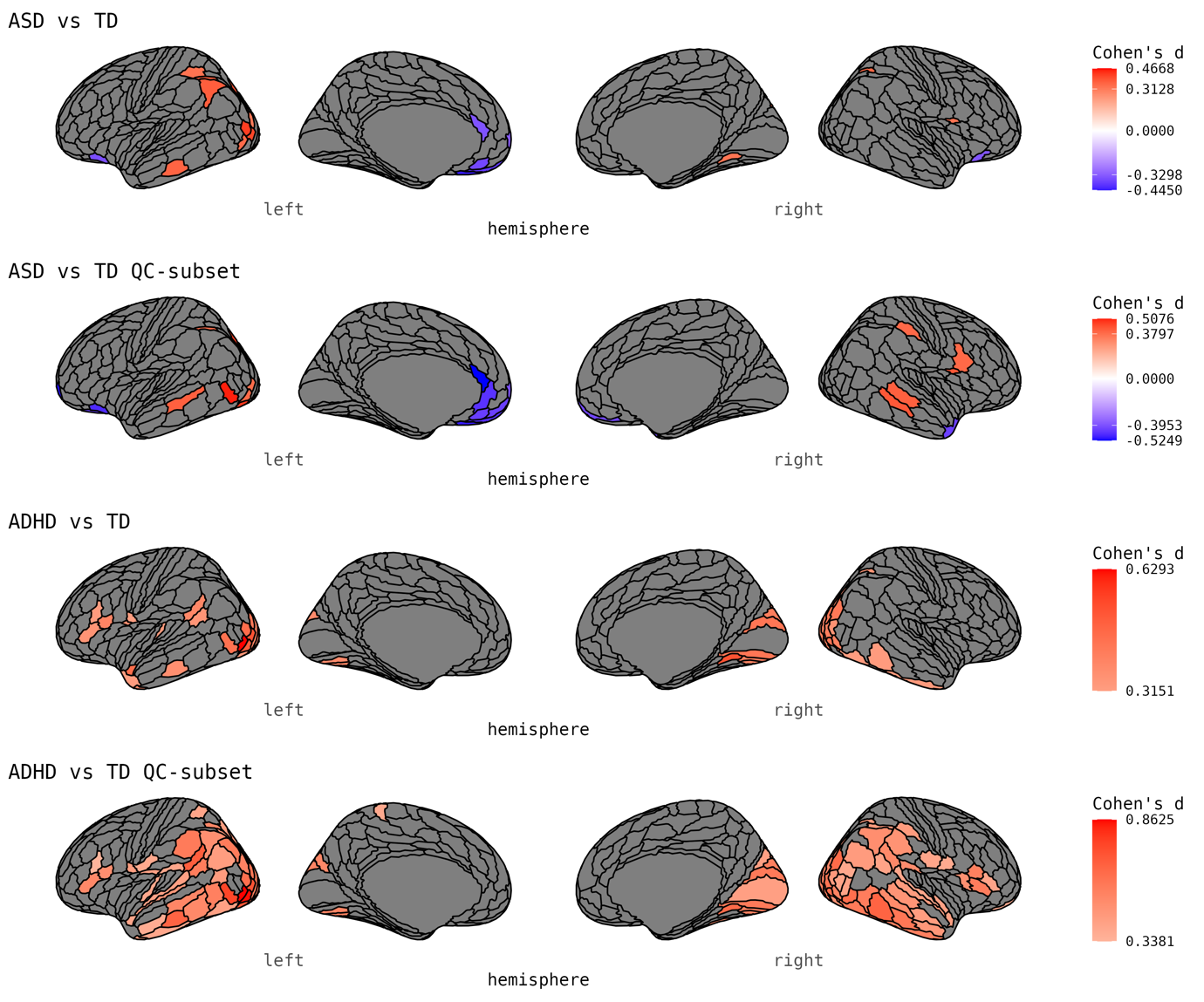

Supplementary Figure 2. Distributions of Cohen’s d of group differences between ASD-, ADHD and TD individuals. The figure shows the distributions of Cohen’s d from the case-control differences in T1w/T2w-ratio. Cohen’s d values are masked by *uncorrected* p-values thresholded at a minimum –logp of 1.3. The abbreviations are as follows: ASD= Autism Spectrum Disorder, ADHD= attention-deficit/hyperactivity disorder, and TD= typically developing individuals.

*Normative modelling*

Normative modelling significantly reduced the associations of age with T1w/T2w-ratio, cortical thickness, and surface area as presented in SI Figure 3

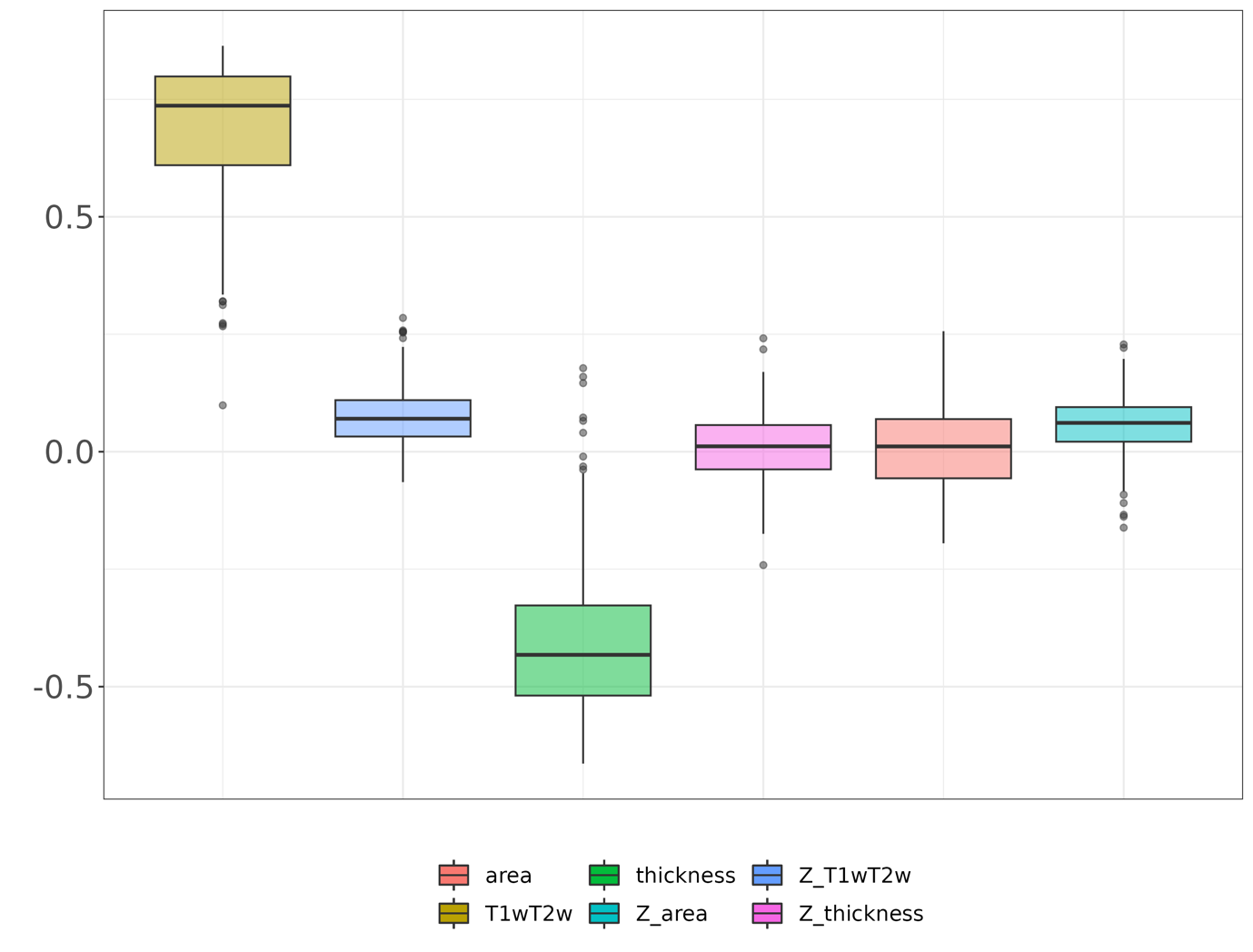

Supplementary Figure 3. Global associations between age and T1w/T2w-ratio, cortical thickness, and surface area pre- and post age-residualization. The figure shows the Pearson’s correlations between original features and age, as well as the z-values outputted from normative modelling and age.

*Unimodal clustering*

Unimodal clustering of T1w/T2w-ratio features showed one relatively large cluster (which further divided at high resolutions), with remaining groupings consisting of single-individual-, or clusters with a very limited number of individuals (SI Figure 4). Further investigations into these clusters showed a similar diagnostic distribution (SI Figure 5), but that individuals appeared to be singled out based on certain characteristics. Either individuals showed a highly local increase in T1w/T2w-ratio, making them an outlier on a single feature, or they had globally increased T1w/T2w-ratio (SI Figure 6), also coupled with a high amount of surface holes (SI Figure 7). T1w/T2w-ratio features that were of particular relevance for parsing are presented in SI Figure 8. Estimators for all tested cluster solutions, are presented in SI Figure 9.

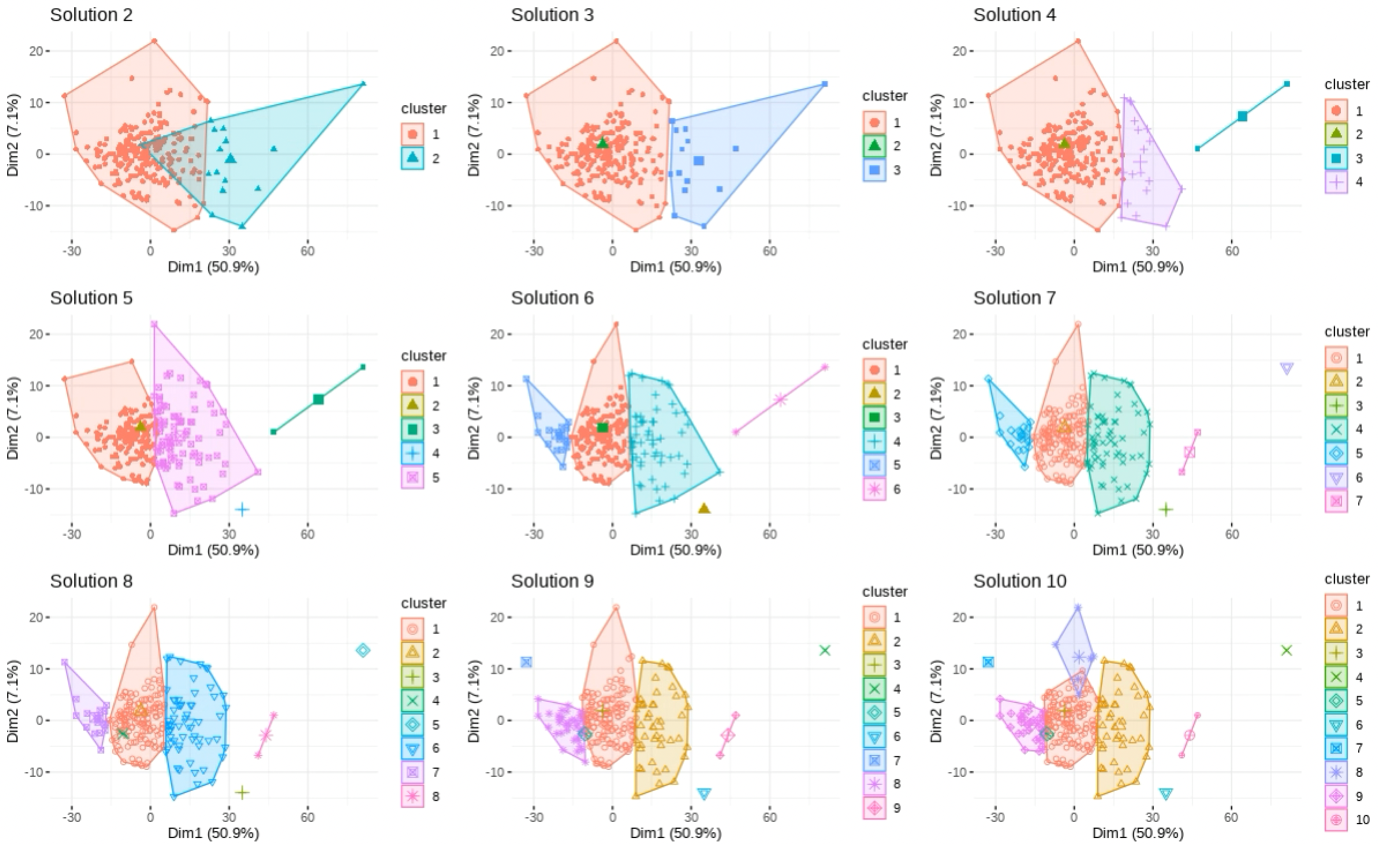

Supplementary Figure 4. Unimodal clustering solutions. The figure illustrates cluster solutions ranging from 2 to 10 from clustering based on T1w/T2w-ratio features. Individuals are plotted according to the first two principal components (Dim1 and Dim2), that explained the majority of the variance in the data. Cluster centroids are depicted as slightly larger shapes.

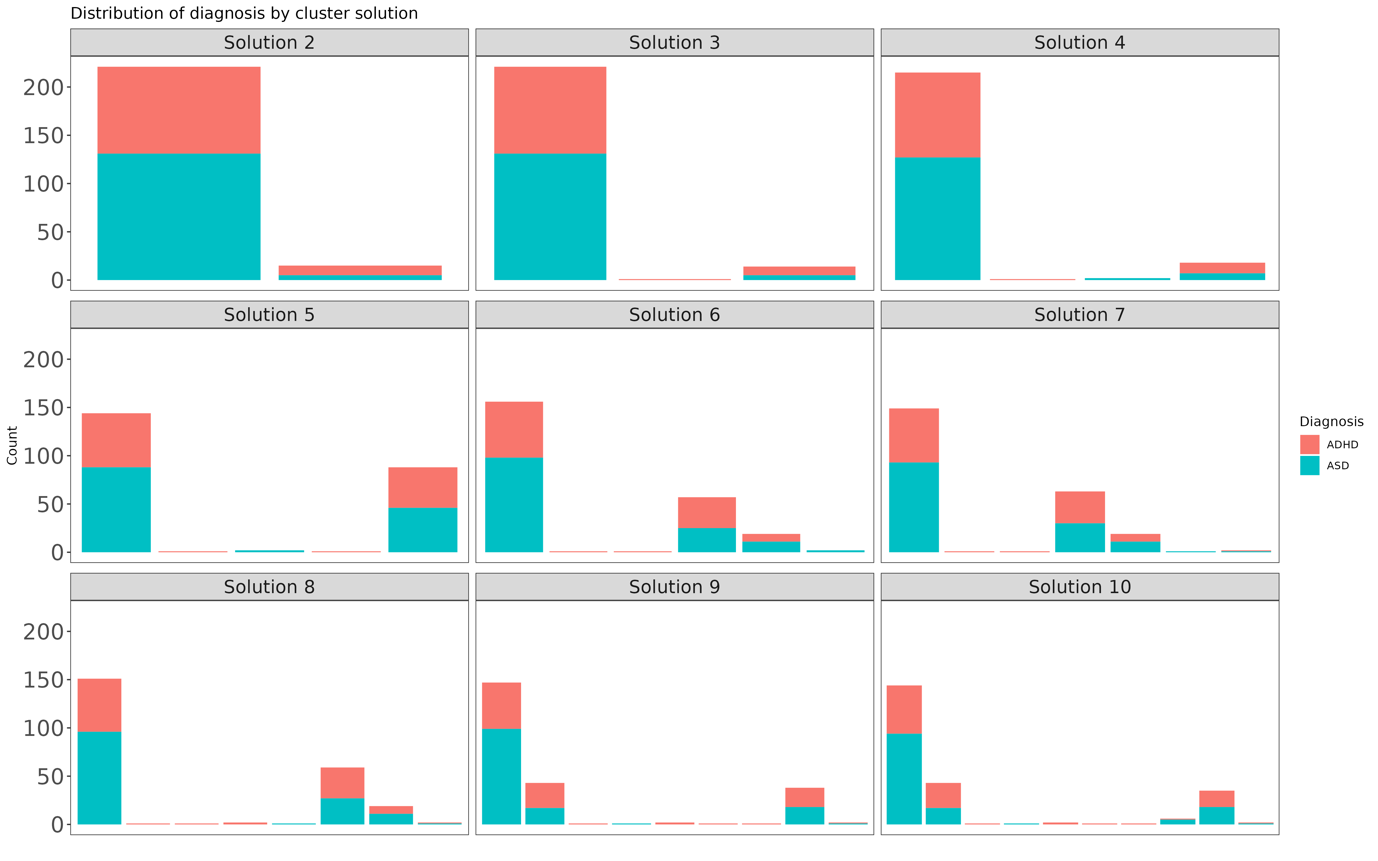

Supplemental Figure 5. Proportion of diagnoses across unimodal clustering solutions. The figure shows bar plots of the proportion of autism spectrum disorder (ASD) and attention deficit hyperactivity disorder (ADHD) across unimodal clustering solutions.

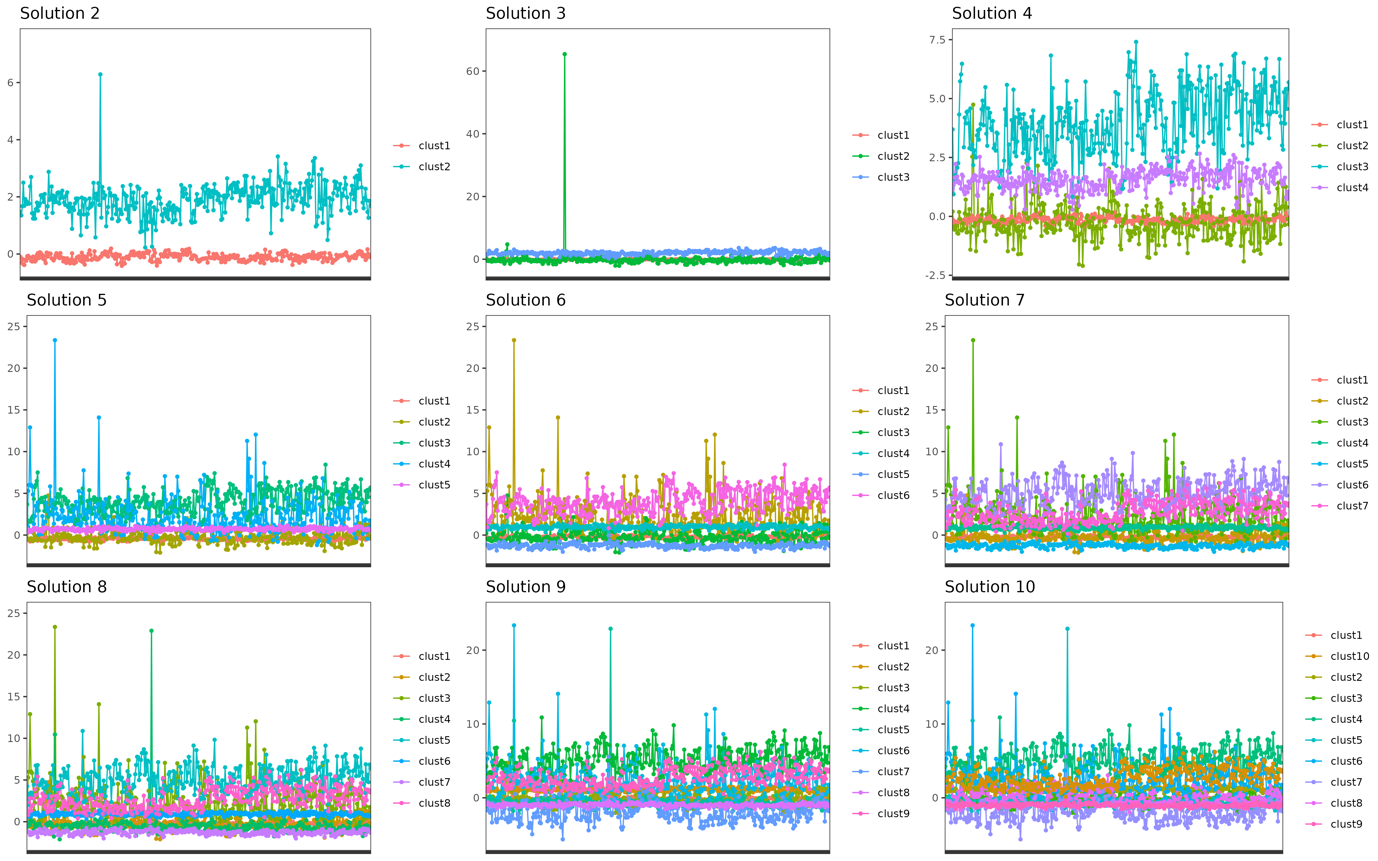

Supplemental Figure 6. Mean T1w/T2w-ratio z-scores across unimodal clustering solutions. The figure illustrates mean T1w/T2w-ratio z-scores for each cluster ("clust"), across different unimodal cluster solutions. Notably, the y-axis for Solution 3 is adjusted to encompass an extreme z-score for a singular feature. Although subsequent cluster divisions also include this outlier, their plots have been set with a lower y-axis threshold to provide a more detailed comparison across solutions.

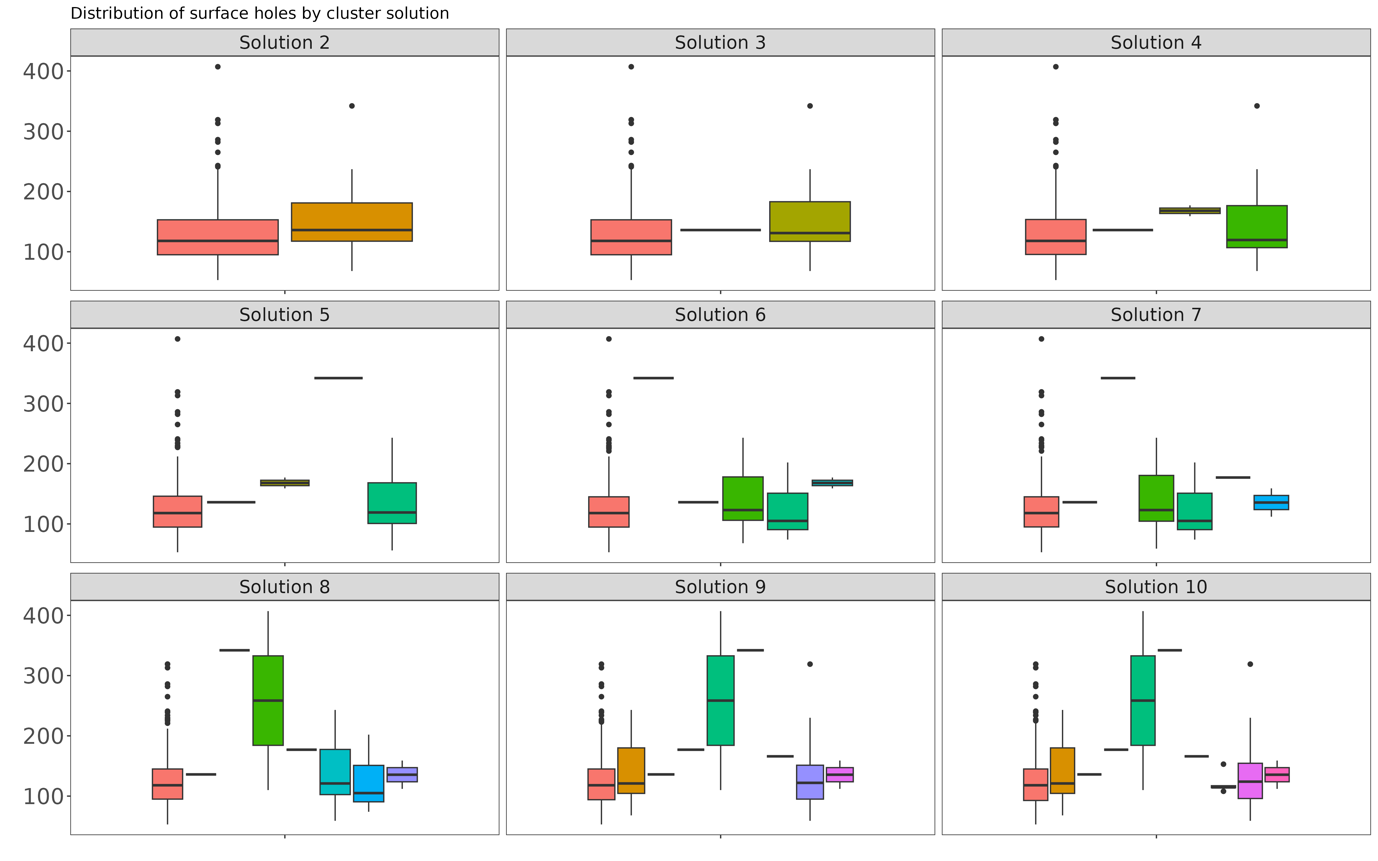

Supplementary Figure 7. Distribution of surface holes across unimodal clustering solutions. The figure shows boxplots of the distribution of number of surface holes within each cluster, separately for each unimodal clustering solution.

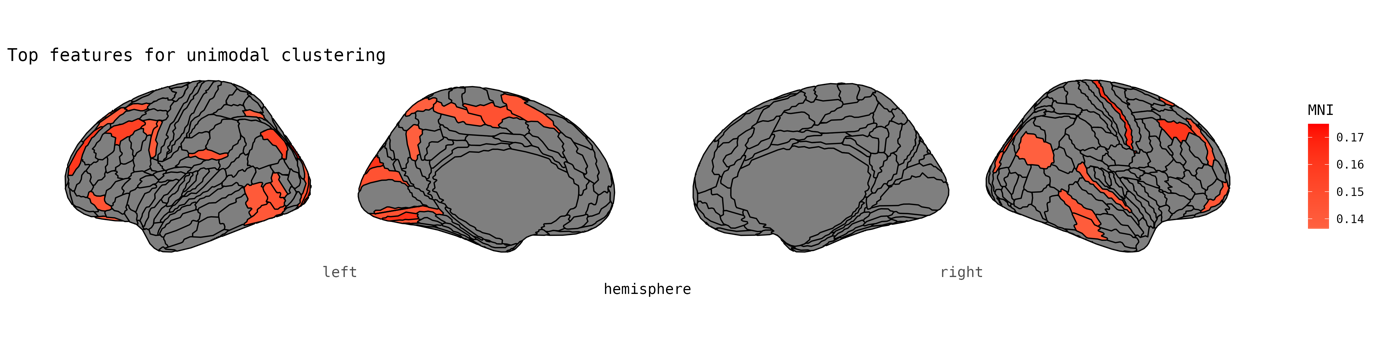

Supplementary Figure 8. The top 10% most central cortical features for unimodal clustering. The figure displays the top 10% most central T1w/T2w-ratio features for clustering, based on the highest ranked normalized mutual information (NMI).

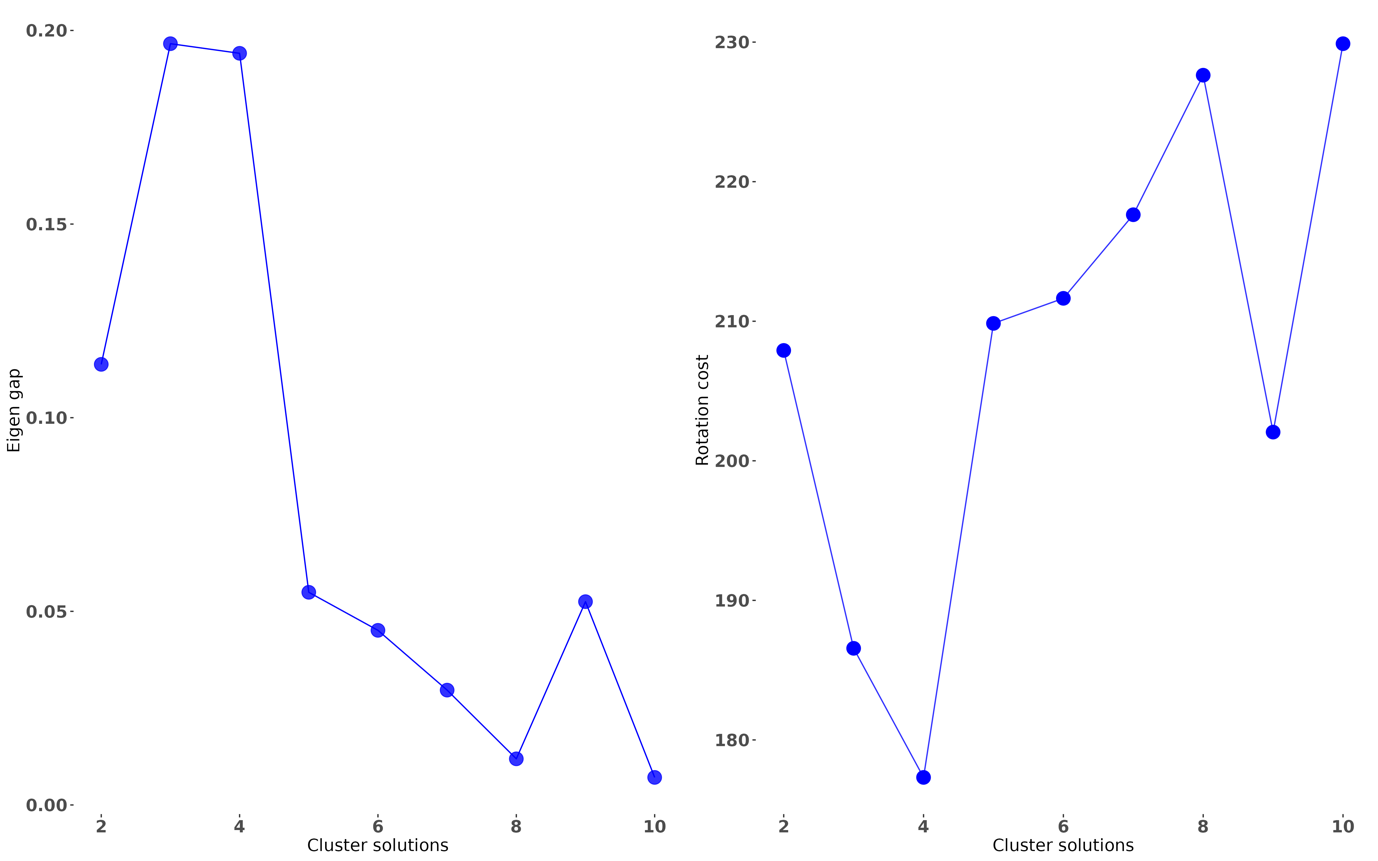

Supplementary Figure 9. Eigen gap and rotation cost of unimodal clustering solutions. The plot shows the eigen gap and rotation cost for unimodal clustering solutions from 2-10.

*Multimodal clustering*

Multimodal clustering of all 1080 features from T1w/T2w-ratio, cortical thickness and surface area resulted in groups of quite equal size (SI Figure 10). The distributions of age, sex, clinical diagnosis, amount of surface holes, and brain volumes are presented in SI Figures 11-15.

Estimators revealed that partitioning into 3 subgroups (eigen gap= 0.045, rotation cost= 199.812) yielded the most optimal solution, which was followed by partitioning into 2 subgroups (eigen gap= 0.036, rotation cost= 209.348). Estimators for all tested cluster solutions, are presented in SI Figure 16.

In the two-subgroup parsing, individuals within Cluster 1 showing statistically higher IQ scores (t=-2.65, p= 0.009 corrected p= 0.045) as compared to individuals within Cluster 2 after correction for multiple testing, across 5 tests. We observed no statistically significant differences among individuals categorized into two subgroups (see distribution in SI Figure 21) and any of the clinical scores. All t-statistics and corresponding p-values are presented in SI Table 1 and 2.

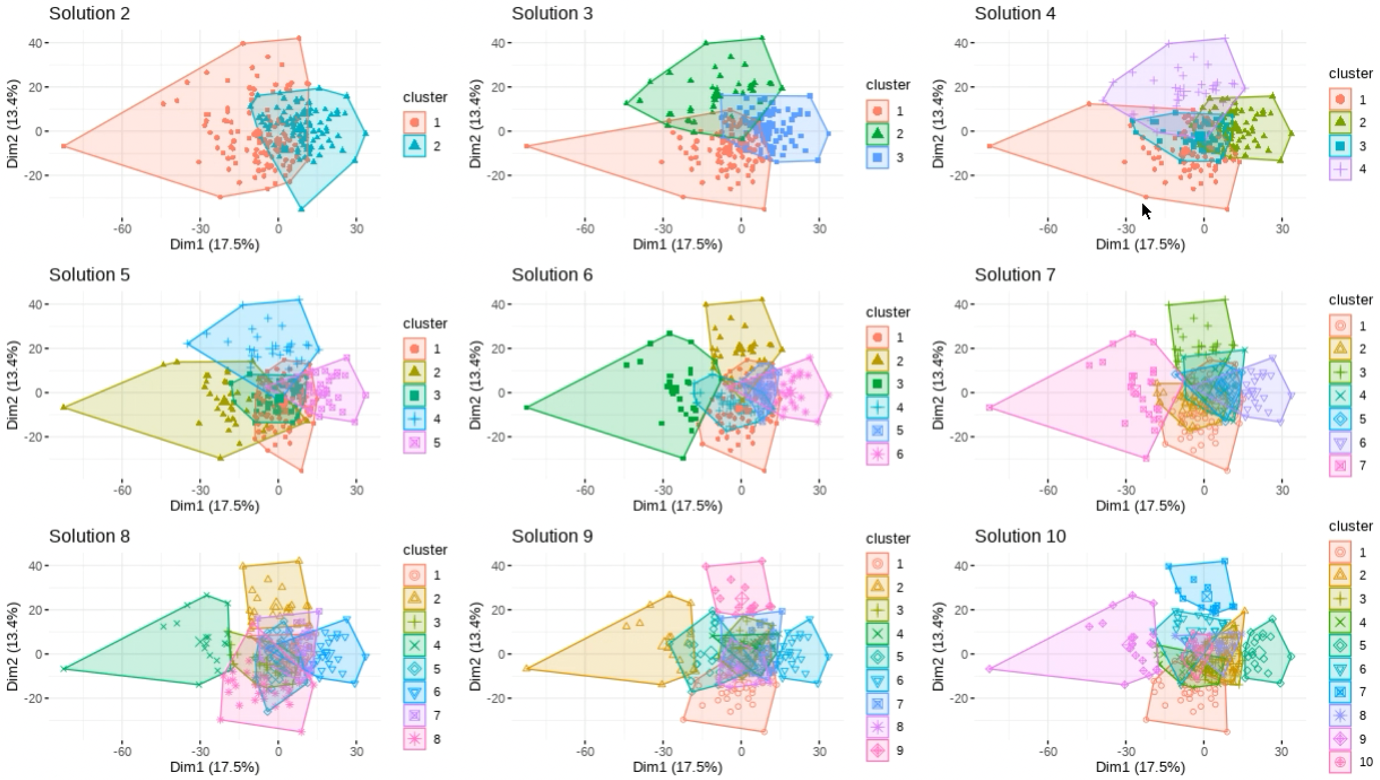

Supplementary Figure 10. Multimodal clustering solutions. The figure illustrates cluster solutions ranging from 2 to 10 from multimodal clustering based on T1w/T2w-ratio, cortical thickness and surface area. Individuals are plotted according to the first two principal components (Dim1 and Dim2), which explained most of the variance in the data. Cluster centroids are depicted as slightly larger shapes.

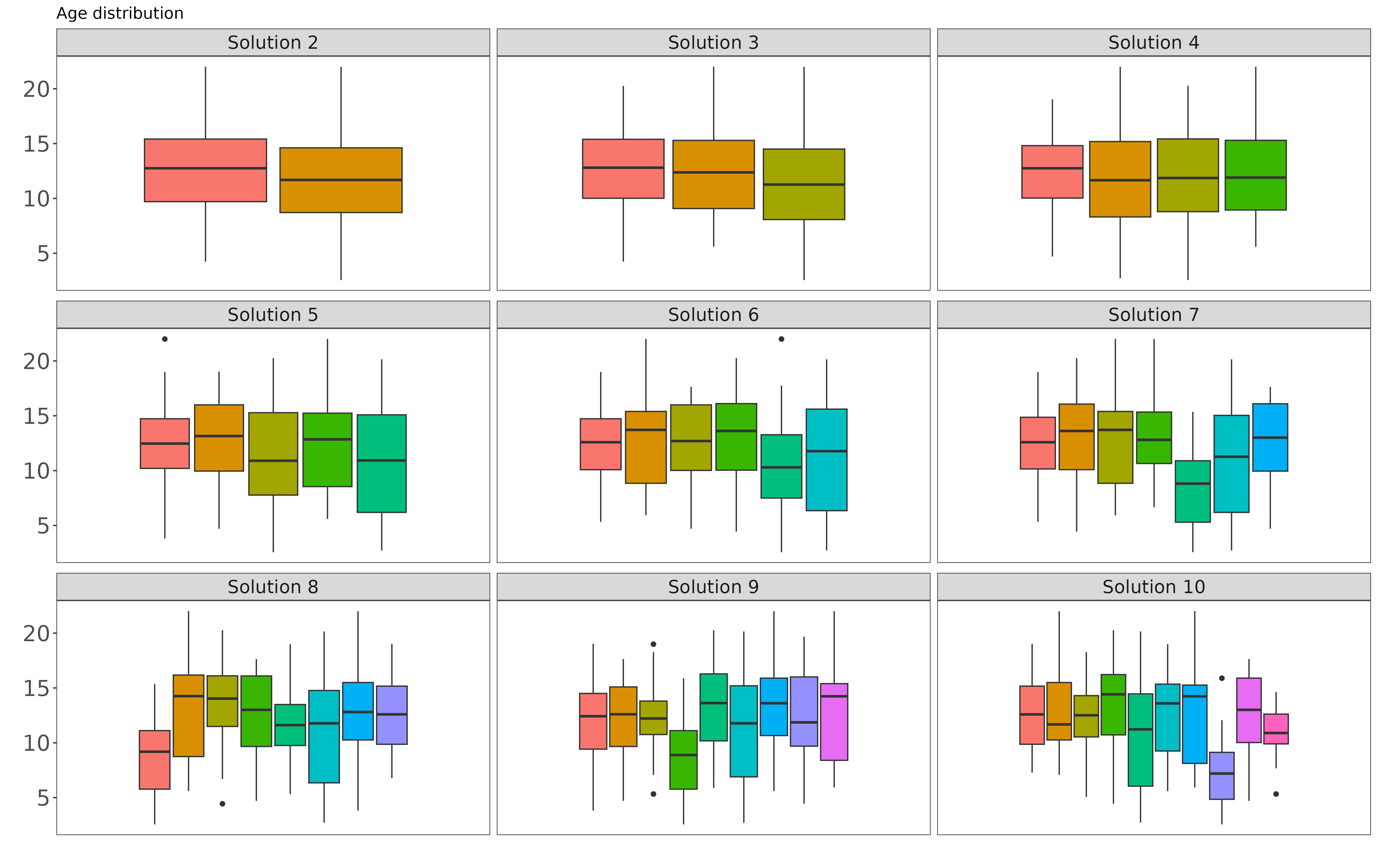

Supplementary Figure 11. Distribution of age across multimodal clustering solutions. The figure shows boxplots of the distribution of age within each cluster, separately for each multimodal clustering solution.

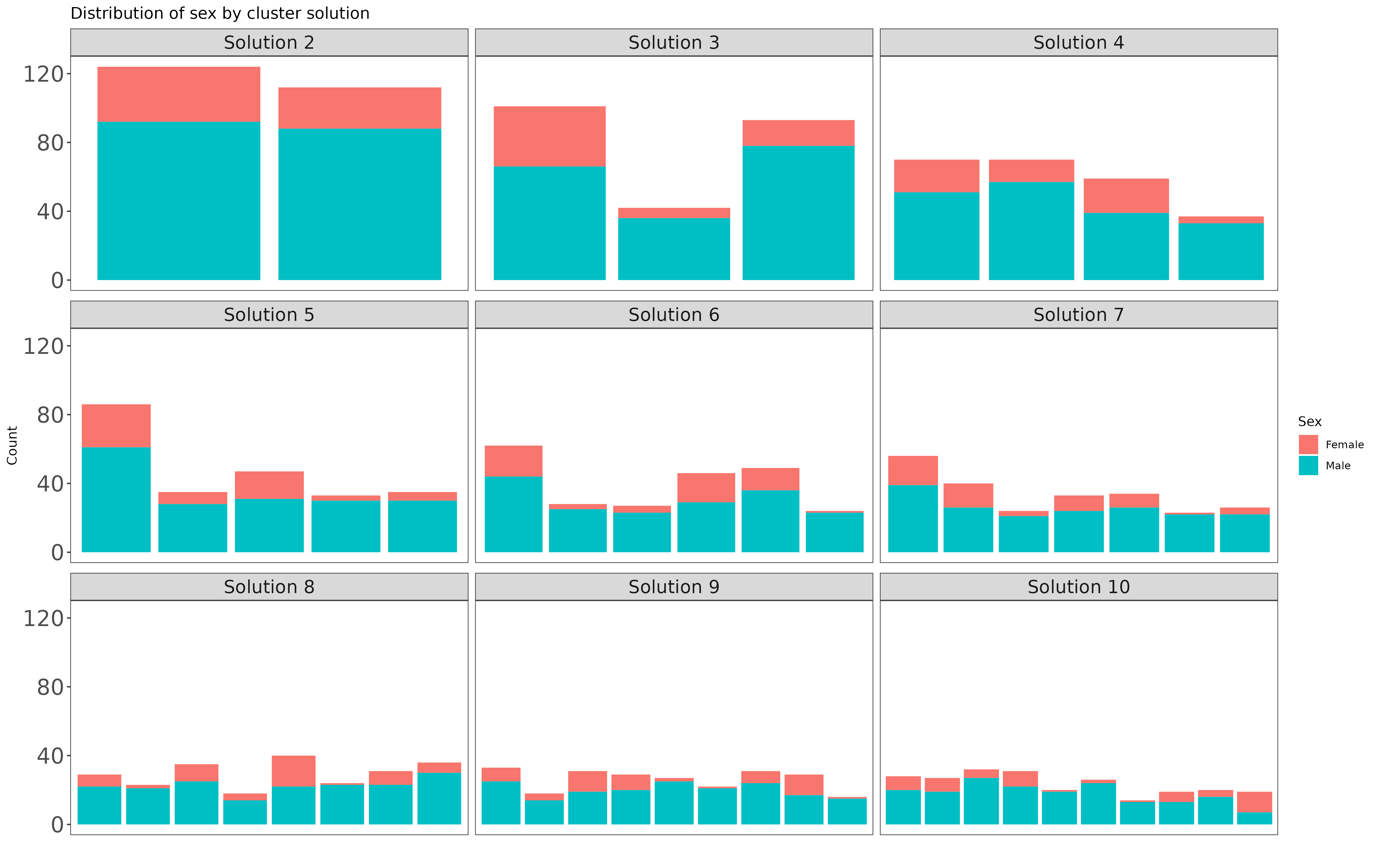

Supplementary Figure 12. Proportion of sex across multimodal clustering solutions. The figure shows bar plots of the proportion of sex across multimodal clustering solutions.

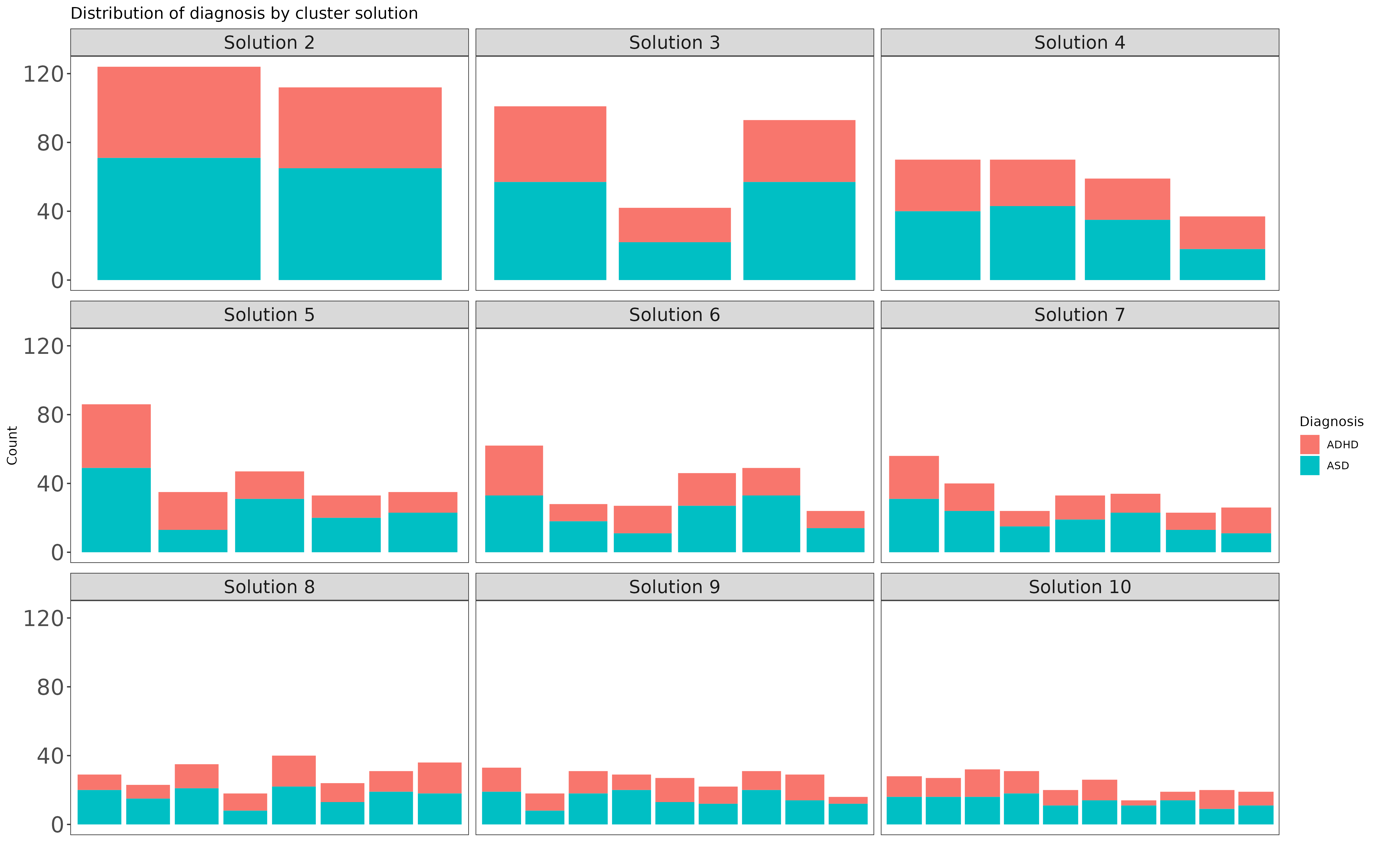

Supplemental Figure 13. Proportion of diagnoses across multimodal clustering solutions. The figure shows bar plots of the proportion of autism spectrum disorder (ASD) and attention deficit hyperactivity disorder (ADHD) across multimodal clustering solutions.

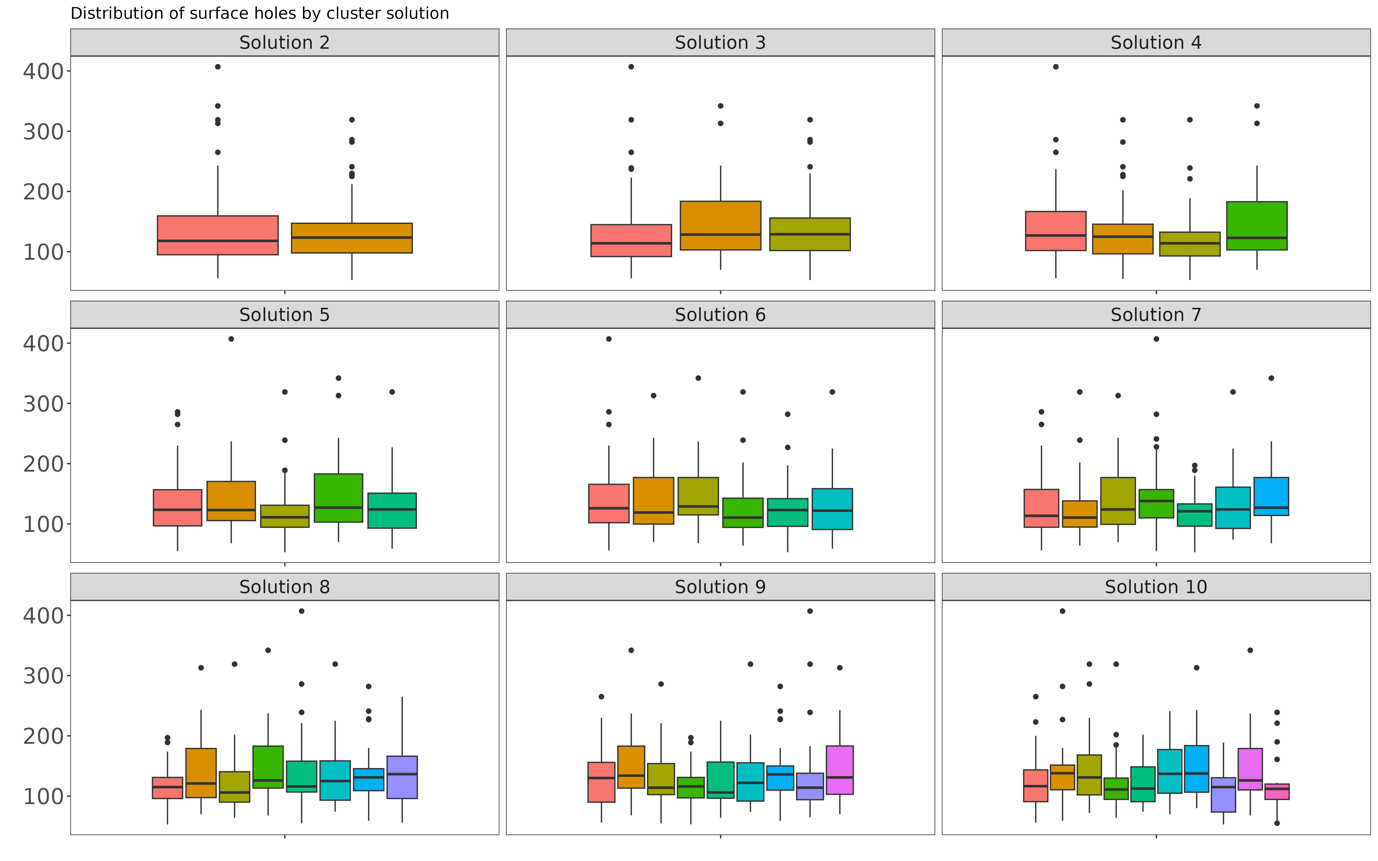

Supplemental Figure 14. Distribution of surface holes across multimodal clustering solutions. The figure shows boxplots of the distribution of surface holes within each cluster, separately for each multimodal clustering solution.

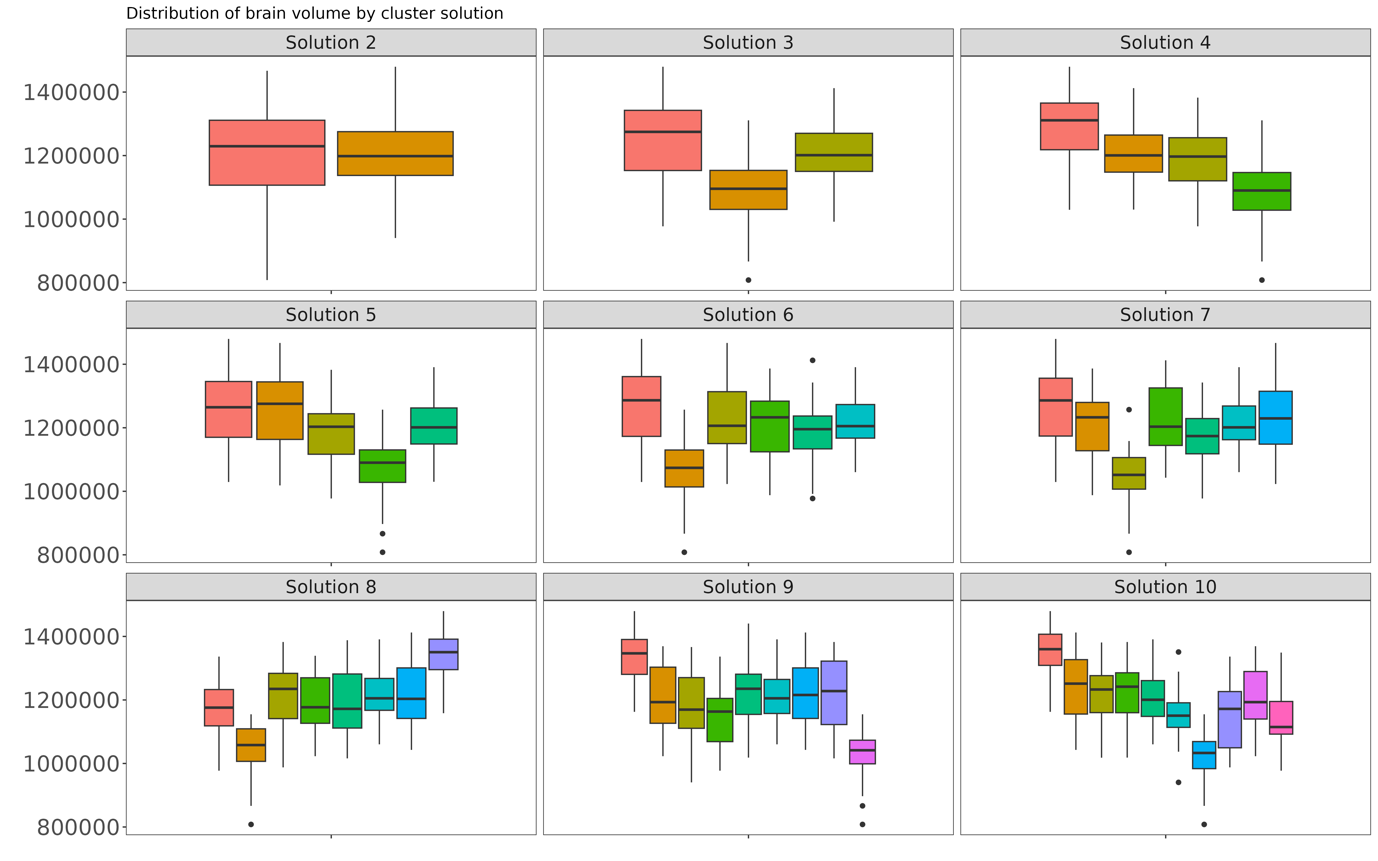

Supplementary Figure 15. Distribution of brain volume across multimodal clustering solutions. The figure shows boxplots of the distribution of brain volume within each cluster, separately for each multimodal clustering solution.

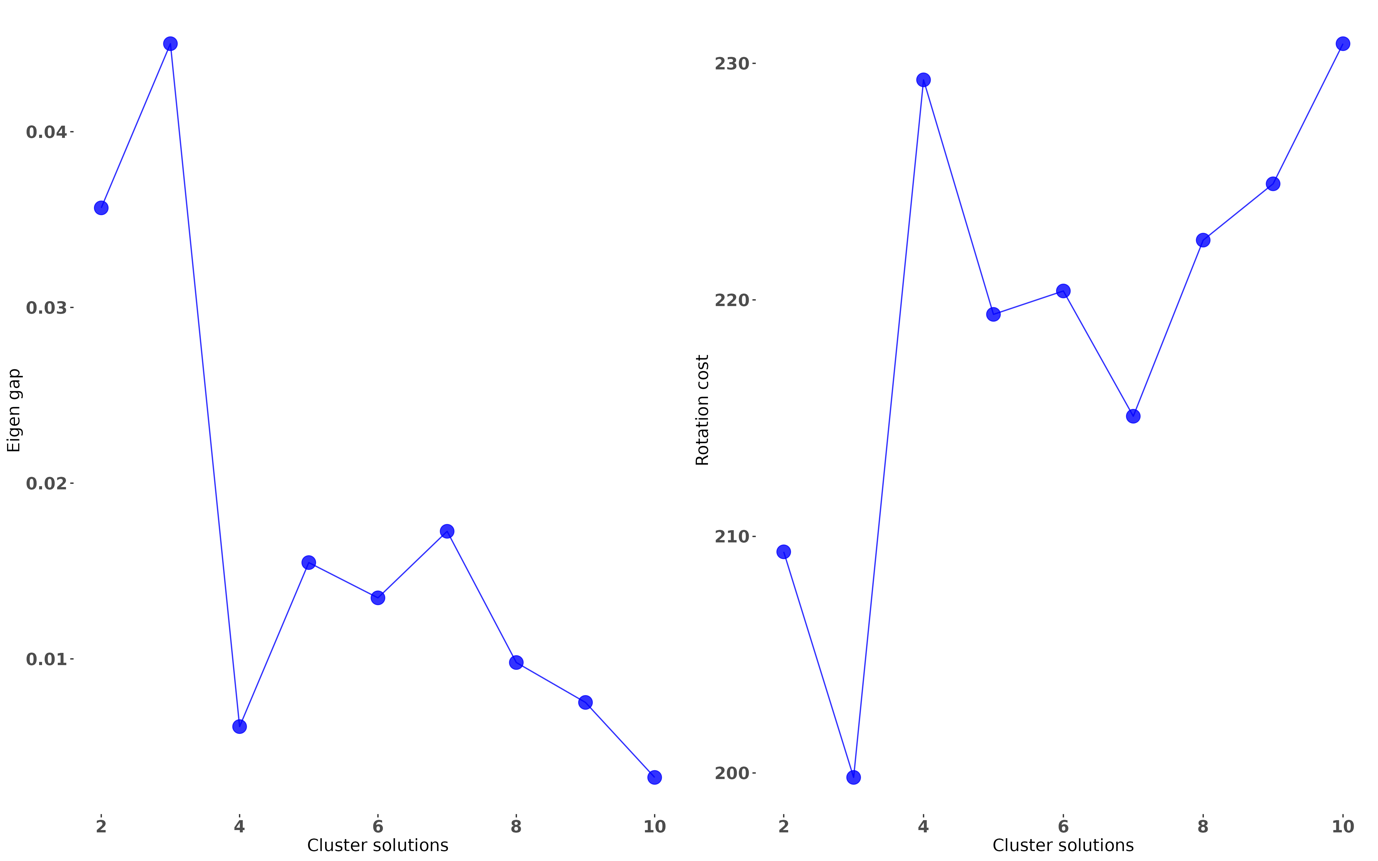

Supplementary Figure 16. Eigen gap and rotation cost of multimodal clustering solutions. The plot shows the eigen gap and rotation cost for multimodal clustering solutions from 2-10.

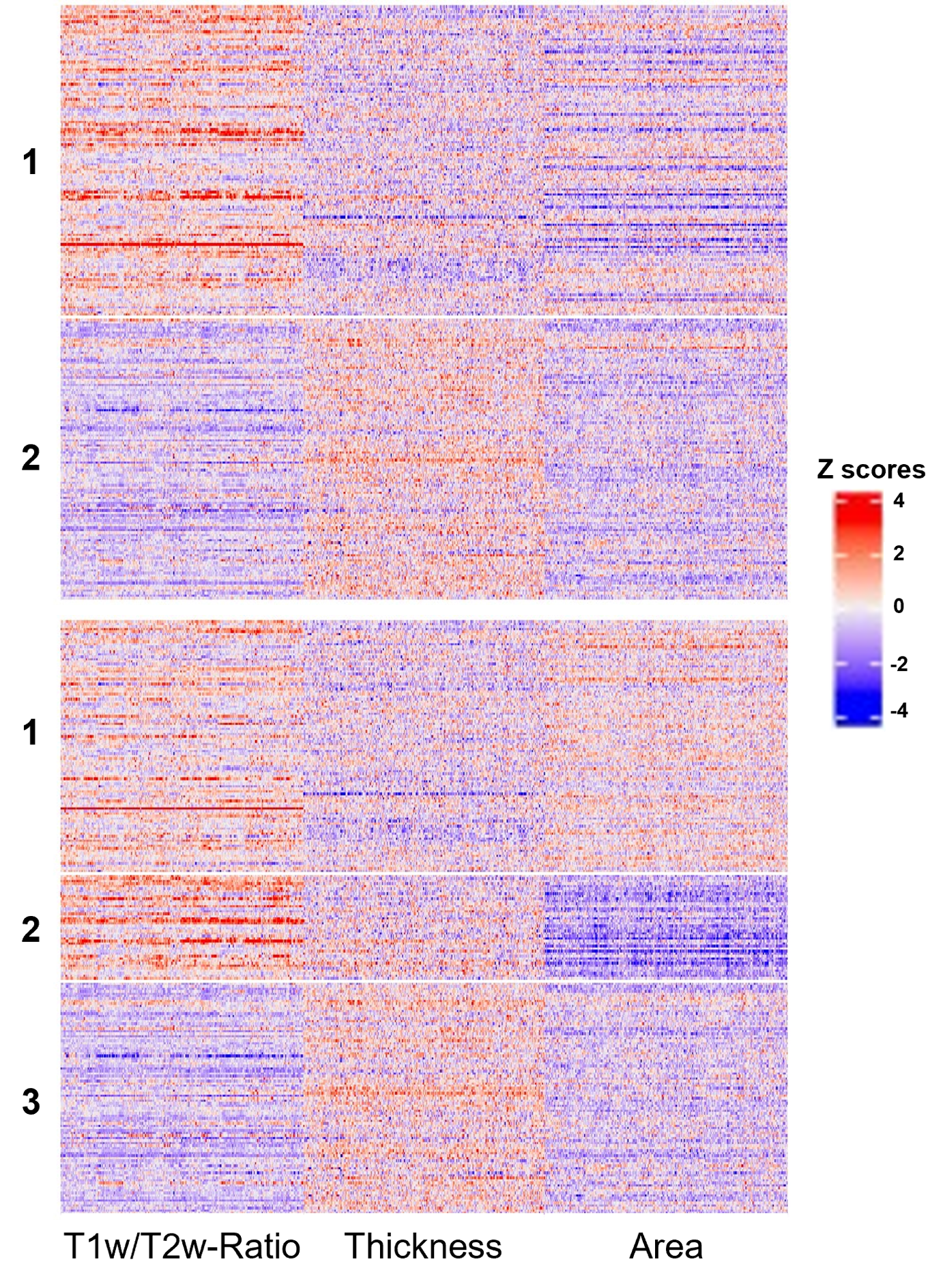
 Supplemental Figure 17. Cortical feature Z-score heatmaps by cluster-membership. The heat maps display z-scores for T1w/T2w ratio, cortical thickness (Thickness) and surface area (Area) across the x-axis, with each row presenting an individual and grouped according to their cluster membership.

*Multimodal clustering in subsets with narrower age ranges*

Boxplots indicated that while some age differences between Cluster 1 and Cluster 3 remained in the n = 180 sample, most of the residual age effects appear to be mitigated within the narrower 8–15 age range (n = 127) (Supplementary Figure 18). Clustering solutions appeared largely consistent across the three sample sizes, indicating good correspondence even when age effects were further minimized (Supplementary Figure 19).

Re-running multimodal clustering with two narrower age range subsets produced cortical patterns largely consistent with the original findings (Supplementary Figure 20). The original Cluster 1, characterized by a slightly higher T1w/T2w ratio, thinner cortex, and somewhat larger surface area, corresponded to Cluster 2 in both re-runs, though with slightly fewer individuals. Similarly, the original Cluster 2, defined by a higher T1w/T2w ratio and much smaller surface area, corresponded to Cluster 3 in the n = 180 subset and Cluster 1 in the n = 127 subset. Lastly, the original Cluster 3, displaying the opposite pattern of Cluster 1 (lower T1w/T2w ratio, thicker cortex, and smaller surface area), corresponded to Cluster 1 in the n = 180 subset and Cluster 3 in the n = 127 subset.

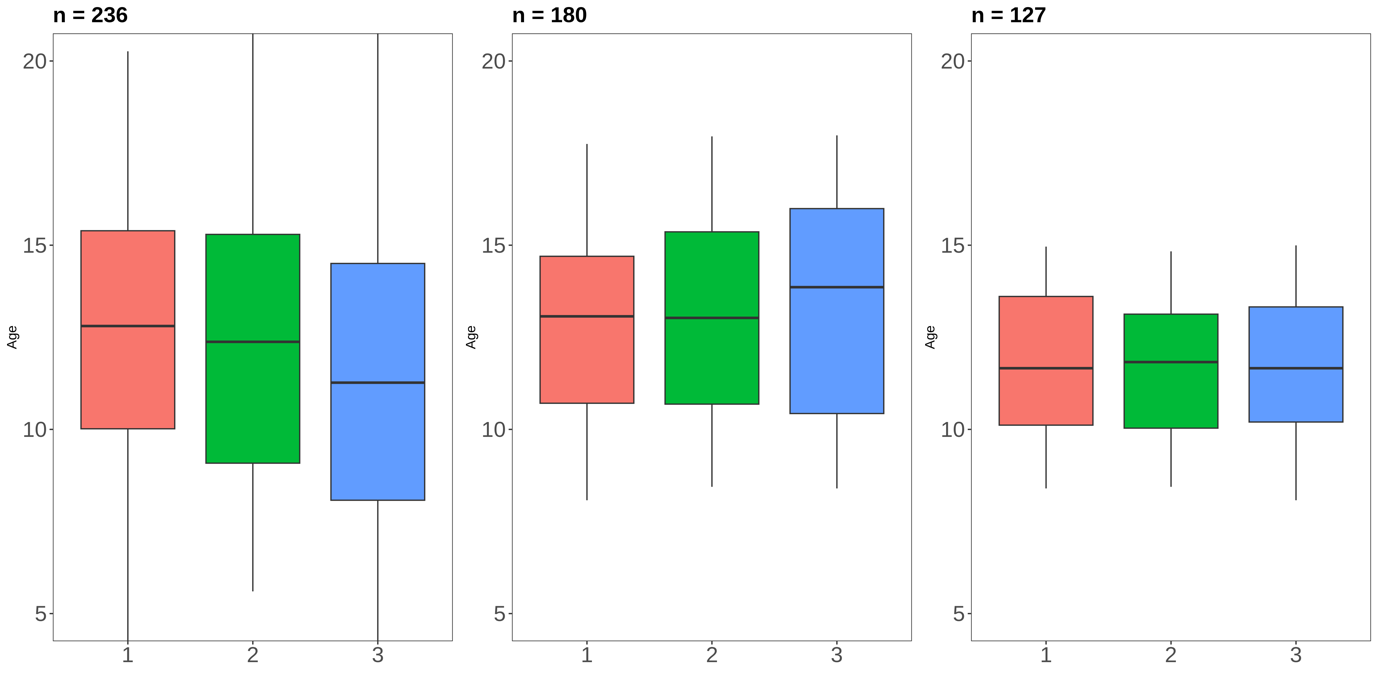

Supplementary Figure 18. Boxplots illustrating the range, median, and interquartile spread of age distributions across cluster solution 3. The leftmost panel shows the age distribution for the original clustering (n = 236). The middle panel represents the narrower age range (8–18 years; n = 180). The rightmost panel displays the further narrowed age range (8–15 years; n = 127).

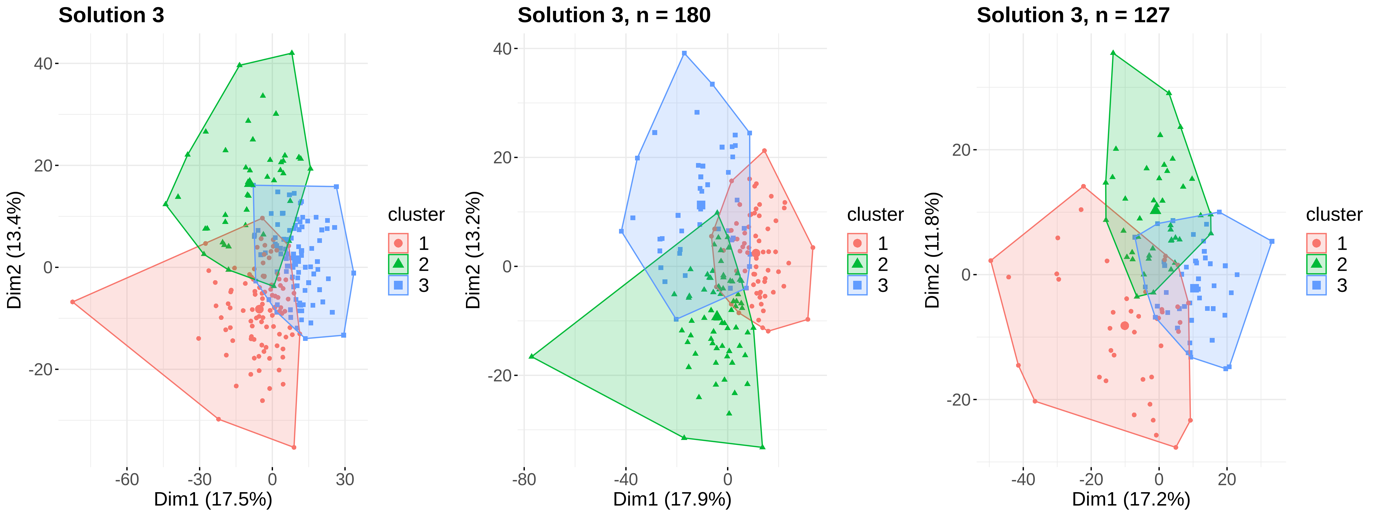

Supplementary Figure 19. The figure illustrates cluster solutions from multimodal clustering based on T1w/T2w ratio, cortical thickness, and surface area. Cluster solutions are visualized across different sample sizes: the leftmost panel represents the full sample size (n = 236), while the middle and rightmost panel represent the intermediate sample size (n = 180) and smallest sample (n = 127) respectively. Individuals are plotted according to the first two principal components (Dim1 and Dim2), which explain the majority of the variance in the data. Cluster centroids are shown as slightly larger markers, and convex hulls delineate the spatial boundaries of each cluster in the multidimensional space.

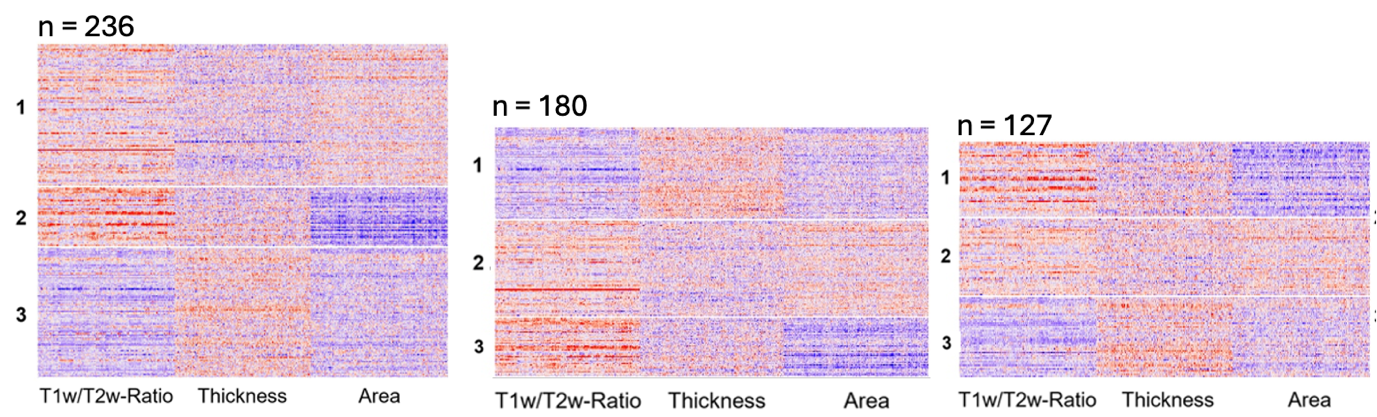

Supplementary Figure 20: Cortical feature Z-score heatmaps by cluster-membership. The heat maps display z-scores for T1w/T2w ratio, cortical thickness (Thickness) and surface area (Area) across the x-axis, with each row presenting an individual and grouped according to their cluster membership. Heat maps are visualized across different sample sizes: the left panel represents the full sample size (n = 236), while the middle and right panels represent the intermediate sample size (n = 180) and smallest sample (n = 127) respectively.

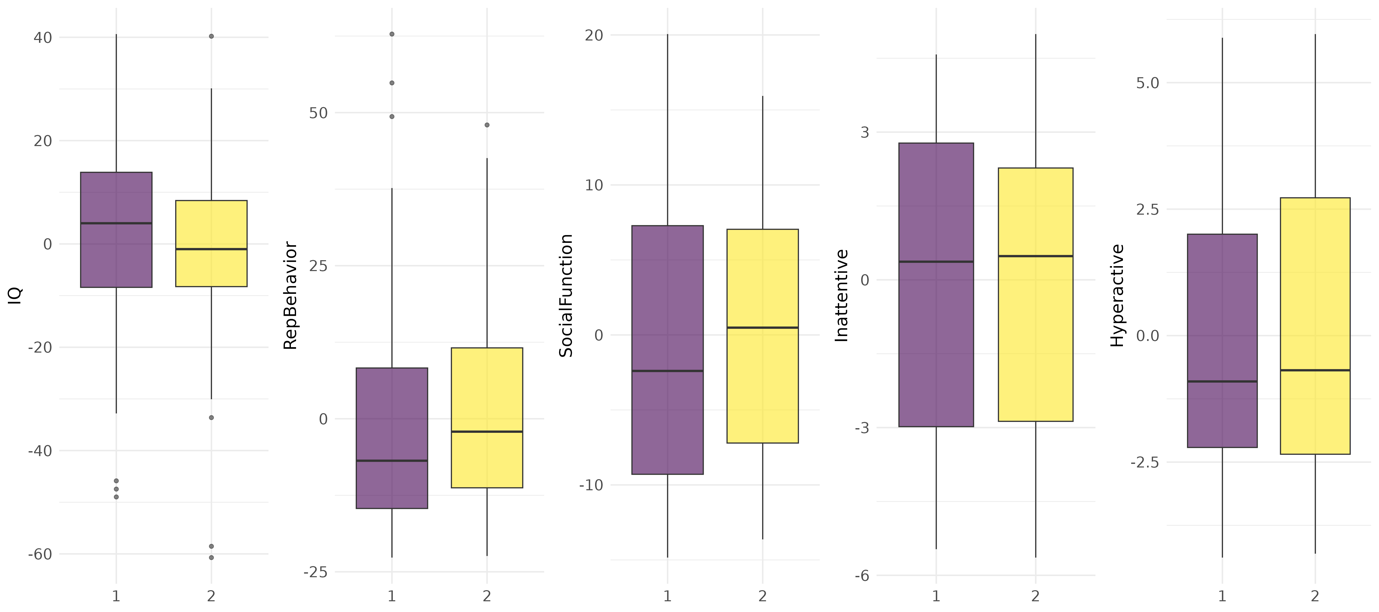

Supplementary Figure 21. Distribution of clinical and cognitive scores for multimodal clustering solution 2. The figure shows boxplots illustrating the distribution of IQ, repetitive behaviors (RepBehavior), social functioning (SocialFunction), inattentive and hyperactive scores after age-residualization. Individuals with missing scores: IQ n= 69, Social Communication Questionnaire (SCQ) n = 72, Repetitive Behaviors Scale – Revised (RBS-R) n = 75, and Strengths and Weaknesses of ADHD-symptoms and Normal-behavior (SWAN) n = 79, are not included in the plots.

| Variable | t-values | p-values | corrected p-values |
| --- | --- | --- | --- |
| IQ | -2.65 | 0.009* | 0.045* |
| RepBehavior | 1.10 | 0.271 | 0.452 |
| SocialFunction | 1.74 | 0.083 | 0.208 |
| Inattentive | 0.36 | 0.721 | 0.901 |
| Hyperactive | -0.09 | 0.930 | 0.930 |

Supplementary Table 1. Clinical and cognitive tests for the 2-cluster solution. The table displays t-statistics and coresponding uncorrected (p-values) and corrected p-values from bootstrapping of quantile regression of the median. An asterisk (*) indicates a p-value <= 0.05.

| Variable and test | t-value | p-value | Corrected p-value |
| --- | --- | --- | --- |
| IQ Cluster 1-2 | -1.10 | 0.271 | 0.581 |
| IQ Cluster 1-3 | -2.82 | 0.005* | 0.075 |
| IQ Cluster 2-3 | 0.17 | 0.864 | 0.948 |
| Repetitive Behaviors Cluster 1-2 | 2.12 | 0.036* | 0.180 |
| Repetitive Behaviors Cluster 1-3 | 1.70 | 0.091 | 0.273 |
| Repetitive Behaviors Cluster 2-3 | -0.62 | 0.536 | 0.815 |
| Social Function Cluster 1-2 | 0.73 | 0.466 | 0.815 |
| Social Function Cluster 1-3 | 1.21 | 0.226 | 0.565 |
| Social Function Cluster 2-3 | 0.61 | 0.543 | 0.815 |
| Inattentiveness Cluster 1-2 | 2.12 | 0.036* | 0.180 |
| Inattentiveness Cluster 1-3 | 1.88 | 0.061 | 0.229 |
| Inattentiveness Cluster 2-3 | 0.08 | 0.936 | 0.948 |
| Hyperactive-Impulsive Cluster 1-2 | 0.13 | 0.893 | 0.948 |
| Hyperactive-Impulsive Cluster 1-3 | 0.34 | 0.736 | 0.948 |
| Hyperactive-Impulsive Cluster 2-3 | -0.07 | 0.948 | 0.948 |

Supplementary Table 2. Clinical and cognitive test for the 3-cluster solution. The table displays t-statistics and coresponding uncorrected (p-values) and corrected p-values from bootstrapping of quantile regression of the median. An astrix (* )indicates a p-value <= 0.05.
